# Mapping behavioural, cognitive and affective transdiagnostic dimensions in frontotemporal dementia

**DOI:** 10.1101/2021.10.29.21265655

**Authors:** Siddharth Ramanan, Hashim El-Omar, Daniel Roquet, Rebekah M. Ahmed, John R. Hodges, Olivier Piguet, Matthew A. Lambon Ralph, Muireann Irish

## Abstract

Two common clinical variants of frontotemporal dementia (FTD) are the behavioural variant (bvFTD) presenting with behavioural and personality changes attributable to prefrontal atrophy, and semantic dementia (SD) displaying early semantic dysfunction primarily due to anterior temporal degeneration. Despite representing independent diagnostic entities, mounting evidence indicates overlapping cognitive-behavioural profiles in these syndromes, particularly with disease progression. Why such overlap occurs remains unclear. Understanding the nature of this overlap, however, is essential to improve early diagnosis, characterisation, and management of those affected. Here, we explored common cognitive-behavioural and neural mechanisms contributing to heterogeneous FTD presentations, irrespective of clinical diagnosis. This transdiagnostic approach allowed us to ascertain whether symptoms not currently considered core to these two syndromes are present in a significant proportion of cases and explore the neural basis of clinical heterogeneity. Sixty-two FTD patients (31 bvFTD, 31 SD) underwent comprehensive neuropsychological, behavioural, and structural neuroimaging assessments. Orthogonally-rotated principal component analysis of neuropsychological and behavioural data uncovered eight statistically independent factors explaining the majority of cognitive-behavioural performance variation in bvFTD and SD. These factors included Behavioural changes, Semantic dysfunction, General Cognition, Executive function, Initiation, Disinhibition, Visuospatial function, and Affective changes. Marked individual-level overlap between bvFTD and SD was evident on the Behavioural changes, General Cognition, Initiation, Disinhibition, and Affective changes factors. Compared to bvFTD, SD patients displayed disproportionate impairment on the Semantic dysfunction factor, whereas greater impairment on Executive and Visuospatial function factors was noted in bvFTD. Both patient groups showed comparable magnitude of atrophy to frontal regions, whereas severe temporal lobe atrophy was characteristic of SD. Whole-brain voxel-based morphometry correlations with emergent factors revealed associations between fronto-insular and striatal grey matter changes with Behavioural, Executive, and Initiation factor performance, bilateral temporal atrophy with Semantic dysfunction factor scores, parietal-subcortical regions with General Cognitive performance, and ventral temporal atrophy associated with Visuospatial factor scores. Together, these findings indicate that cognitive-behavioural overlap (i) occurs systematically in FTD, (ii) varies in a graded manner between individuals, and (iii) is associated with degeneration of different neural systems. Our findings suggest that phenotypic heterogeneity in FTD syndromes can be captured along continuous, multidimensional spectra of cognitive-behavioural changes. This has implications for the diagnosis of both syndromes amidst overlapping features as well as the design of symptomatic treatments applicable to multiple syndromes.

## Introduction

Frontotemporal dementia (FTD) refers to a group of rare, young-onset, progressive neurodegenerative brain disorders, primarily affecting the frontal and/or temporal lobes.^1^ Three clinical variants have been defined: a behavioural variant (bvFTD), a semantic variant (here, referred to as semantic dementia or SD), both of which form the focus of the current study, and a non-fluent aphasic variant. Briefly, bvFTD presents with marked changes in behaviour, personality, executive and social cognition, in the context of marked prefrontal and insular degeneration.^2^ In contrast, SD is characterised by profound anomia and comprehension difficulties reflecting trans-modal, trans-category loss of semantic knowledge arising from degeneration of bilateral ventral and anterior temporal lobes (ATL).^3-5^ While most SD patients present with left-predominant ATL degeneration (SD-Left), a right-lateralized pattern (SD-Right) has also been described, whereby atrophy initially targets the right hemisphere before producing a bilateral profile of ATL degeneration. These patients typically present with face processing disturbances, socio-emotional dysfunction, behavioural changes, and loss of insight, in addition to semantic impairments.^6-8^

Clinically, bvFTD and SD are considered as separate diagnostic entities, however, mounting evidence suggests the need to rethink traditional phenotypic boundaries between these syndromes,^9^ especially in terms of cognitive and behavioural performance.^8,10-12^ For example, semantic and language changes also occur in bvFTD^13,14^ and are detectable in early disease stages.^10^ Likewise, SD patients (especially SD-Right) present with considerable socioemotional and behavioural disturbances^7,15-19^ often of the same magnitude as bvFTD, and which intensify with disease progression. Estimating the frequency of heterogeneous symptoms from clinical reports, Coyle-Gilchrist^20^ found ∼74% of their referred bvFTD patients (*N*=31/42) to show non-specific language difficulties, whereas ∼96% of their SD cohort (*N*=22/23) displayed behavioural changes. Emerging evidence also suggests considerable overlap between bvFTD and SD on executive, attentional and visuospatial performance.^16^ Currently, it remains unclear why distinct clinical syndromes display overlapping symptoms. Two proposals relevant to FTD are that (i) emergent heterogeneity relates closely to advancing disease severity, and (ii) individuals showing a combination of bvFTD and SD symptoms (with additional prosopagnosia and visuospatial dysfunction) represent a clinically distinct entity such as the ‘right temporal variant of FTD’.^11,21-23^ Both proposals, however, cannot account for ‘atypical’ symptoms in individuals early in the disease trajectory, in patients whose clinical profiles otherwise squarely meet diagnostic criteria for prototypical bvFTD or SD, or for the many patients presenting with ‘mixed’ symptomatology. As these overlaps are not reflected in international diagnostic criteria for bvFTD and SD, quantifying the nature and extent of such variation is important for improved diagnosis and characterisation of both syndromes.

Cognitive-behavioural variations in bvFTD and SD have typically been studied using group comparisons of prevalence/severity of specific deficits. This approach is limited in its capacity to capture individual-level variability, subtle overlaps, and fuzzy between-group boundaries between both syndromes. Instead, transdiagnostic approaches that capture systematic, non-random variations in cognitive-behavioural symptoms across individuals, irrespective of categorical diagnostic labels, offer greater promise in this regard. By cutting across categorical boundaries, this approach aids examination of prevalence and magnitude of cognitive-behavioural features, and their likely associations with the presenting phenotype.^24^ By moving from a diagnosis-centred to symptom-centred approach, we can accommodate features that occur systematically in some patients (e.g., ‘pure’ presentations of bvFTD and SD) as well as symptoms that are ‘diagnostically atypical’ (e.g., occurring in those with ‘mixed profiles’).

Transdiagnostic approaches have been applied consistently in neuropsychiatry and post-stroke aphasia fields to model neurocognitive mechanisms explaining cognitive, behavioural, and functional breakdowns, irrespective of syndrome-specificity.^25-28^ Recent studies employing these methods in dementia syndromes demonstrate considerable success in explaining symptomatic heterogeneity in terms of coherent variations along orthogonal dimensions of clinical and cognitive changes.^8,12,29-32^ Across syndromes, such phenotypic variations further closely relate to unique patterns of neural network degeneration,^8,12,30^ metabolic brain changes,^33^ and categorically-distinct neuropathological drivers of underlying disease.^34^ Applying transdiagnostic approaches in bvFTD and SD, therefore, holds immense promise in understanding clinical heterogeneity of these syndromes and associated neurocognitive mechanisms.

This study aimed to capture the spectrum of cognitive-behavioural features present across bvFTD and SD, considered as a whole, using data-driven, orthogonally-rotated principal component analysis (PCA). When used in a transdiagnostic manner, this method models statistical co-dependencies within and between bvFTD and SD cognitive-behavioural performance data to output a set of core factors or ‘dimensions’. Dimensions are heavily informed by inter-/intra-group performance variations and reflect the involvement of different underlying cognitive processes explaining symptomatic heterogeneity in these syndromes, free from the constraints of categorical labels. By associating these variations to underlying brain network changes, we can further uncover common neural signatures of shared symptomatology in clinically distinct groups. Together, this approach allows us to move *beyond* thinking of heterogeneous and atypical presentations as descriptive of a specific subgroup or subtype of FTD. Instead, we can capture variations occurring commonly *and* uniquely in both groups, allowing us to position bvFTD and SD patients as varying along a continuous, multidimensional FTD space of cognitive-behavioural changes and associated brain dysfunction.

## Materials and methods

### Participants

Ninety-two participants were recruited through FRONTIER, the frontotemporal dementia research group at the Brain and Mind Centre, The University of Sydney, Australia. Thirty-one patients with a clinical diagnosis of probable bvFTD^35^ and 31 patients with a clinical diagnosis of SD^36^ were included. The SD group was further classified into ‘SD-Left’ (*N*=20) or ‘SD-Right’ (*N*=11) based the magnitude and laterality of ATL and temporopolar atrophy on structural Magnetic Resonance Imaging (MRI) (Supplementary Figure 1). Unless subgroups are explicitly mentioned, we considered the SD group as one heterogenous cohort in the analyses.

Diagnoses were established by consensus among a multidisciplinary team comprising senior neurologists (R.M.A, J.R.H), clinical neuropsychologists, and occupational therapists based on comprehensive clinical, neuropsychological, and structural MRI assessments. Disease severity in all patients was established using the Clinical Dementia Rating–Frontotemporal Lobar Degeneration Sum of Boxes score (CDR-FTLD SoB^37^). Carer ratings on the Cambridge Behavioural Inventory–Revised (CBI-R^38^) and the Neuropsychiatric Inventory (NPI^39^) were used to index frequency and severity of behavioural changes in patients.

Thirty healthy Control participants, comparable to patient groups for sex and education, were selected through the research volunteer panel and local community clubs. All Controls scored zero on the CDR-FTLD SoB measure and 88 or above on the Addenbrooke’s Cognitive Examination–Revised (ACE-R^40^) – a global assessment of cognitive function spanning attention, memory, verbal fluency, language, and visuospatial processing domains. Exclusion criteria for all participants included history of significant head injury, cerebrovascular disease, alcohol and drug abuse, other primary psychiatric, neurological or mood disorders, and limited English proficiency.

All participants or their Person Responsible provided written informed consent in accordance with the Declaration of Helsinki. This study was approved by the South Eastern Sydney Local Health District and The University of New South Wales ethics committees.

### General and targeted neuropsychological assessment

Participants underwent comprehensive neuropsychological testing of memory, language, and executive functioning (description, scoring, and relevant references detailed in Supplementary Methods). Briefly, targeted language assessments of single word naming, comprehension, repetition, semantic association (subtests of the Sydney Language Battery or SYDBAT), as well as auditory attention and working memory (forward and backward scales of the Digit Span Task), and tests of controlled word generation (letter fluency) were included. We also measured performance on visuo-construction abilities and visuospatial recall functions (Rey-Osterrieth Complex Figure or ROCF), executive functions (time difference between parts B and A of Trail Making Test or TMT B-A), as well as an emotion recognition and affect selection task (total score on Facial Affect Selection Task or FAST).

### Behavioural assessments

Clinician-rated changes on agitation, depression, anxiety, apathy, disinhibition, and irritability/lability scales from the NPI were assessed (using frequency*severity scores). In addition, subscales of the CBI-R assessing alterations in activities of everyday living, memory, self-care, abnormal and false beliefs, mood, eating, sleep, stereotypical behaviours, and motivation were employed. Finally, carer burden was assessed using the Zarit Burden Interview.^41^

### Statistical analyses

Behavioural data were analysed using RStudio v4.0.0^42^ and MATLAB (The Mathworks Inc., Natick, MA).

For the PCA, all patients were treated as one heterogeneous FTD cohort. For descriptive analyses, visualisations, and comparisons of PCA scores, however, we retain the clinico-anatomical distinction of SD-Left and SD-Right to aid interpretation.

### Step 1: Characterizing group differences

Chi-square tests were used to investigate group differences for categorical variables (*i*.*e*., sex). For continuous variables, Shapiro-Wilk tests and box-and-whisker plots were first used to determine normality of distribution. When data met normality assumptions, *t*-tests or analyses of variance (ANOVA) were used followed by Sidak post-hoc comparisons. Wilcoxon-Mann-Whitney tests were employed when data violated normality assumptions. For all analyses of group differences, an alpha of *p*≤.05 was employed.

### Step 2: Standardizing scores and imputing missing data

All subsequent analyses were run in the patient cohort only (*N*=62). PCA solutions rely on standardized ‘full’ datasets with no missing variables, therefore, missing data were imputed. The combined patient cohort had 4.17% missing data (Supplementary Figure 2). Available data were converted into percentages following which, missing data were imputed using probabilistic PCA with *k*-fold cross-validation (detailed in Supplementary Methods).

### Step 3: Identifying principal cognitive factors using Principal Component Analysis

To determine principal factors of cognitive and behavioural performance, an omnibus PCA employing orthogonal (varimax) rotation was undertaken. Orthogonal rotations maximize dispersion of loadings between components, allow for little shared variance between emergent components, and facilitate clear behavioural and cognitive interpretations. As per recommended approaches^43^, components with eigenvalues >1.0 were extracted and assigned labels to reflect the majority of variables loading heavily (>|.5|) on each component.

At the outset, we clarify that factor names are simply shorthand for functions assessed by the majority of tests loading onto that particular factor and, by no means, reflect the entirety of cognitive or behavioural processes underpinning performance across all variables that belong to that factor.

Individual scores on each emergent factor were extracted and used as orthogonal covariates in subsequent behavioural and neuroimaging correlation analyses. To understand patient factor performance in relation to Control performance, we further projected the lower bound of normality score (−1.96 standard error of the mean) from Control data into the patient’s PCA space (Supplementary Methods). Group differences between patients on factor scores were examined using ANOVAs with Sidak corrections for *post-hoc* comparisons.

### Image acquisition

Eighty-nine participants (29 bvFTD, 30 SD, and 30 Controls) underwent structural MRI using a 3T Philips MRI scanner with a standard quadrature head coil (eight channels). Whole-brain T_1_-weighted images were acquired using the following parameters: coronal acquisition, matrix 256*256 mm, 200 slices, voxel size=1mm^3^, echo time/repetition=2.6/5.8 msec, flip angle α=8°.

### Voxel-based morphometry analyses

Changes in grey matter intensity between groups were investigated using whole brain Voxel-Based Morphometry (VBM) analyses in FSL (FMRIB Software Library: https://fsl.fmrib.ox.ac.uk/fsl/fslwiki). Pre-processing included brain extraction^44^, tissue segmentation^45^, and alignment of segmented images to Montreal Neurological Institute (MNI) standard space using non-linear re-registration^46,47^. Full details of pre-processing steps are detailed in Supplementary Methods.

### Whole brain changes in grey matter intensity

Whole-brain voxel-wise differences in grey matter intensity between bvFTD, SD, and Control groups were examined using independent *t*-tests with age included as a nuisance variable. Clusters were extracted using the Threshold-Free Cluster Enhancement method using a threshold of *p*<.01 corrected for Family-Wise Error (FWE) with a cluster threshold of 100 spatially contiguous voxels.

### Inter-subject variance in magnitude and asymmetry of atrophy in disease-specific epicentres

Prior to decomposing cognitive heterogeneity in FTD patients, it is essential to quantify variations in prefrontal and temporal integrity at the individual level, allowing us to position each patient into a continuous frontotemporal atrophy space. Towards this, we calculated the magnitude and asymmetry of atrophy to key frontal and temporal regions that represent potential epicentres of atrophy in bvFTD and SD, respectively.^8,48,49^ We first selected four regions-of-interest (ROI) in prefrontal (left and right orbitofrontal cortex [OFC]), anterior insula, and temporal cortices (left and right temporal pole and ATL) from established atlases, based on *a priori* knowledge of atrophy epicentres in bvFTD and SD (Supplementary Methods). The ATL masks excluded the temporal poles, so when used in conjunction with temporal pole masks, it allowed us to capture gradation of atrophy along the longitudinal axis of the temporal neocortex (Supplementary Figure 3). For all patients, mean intensity values for each ROI were extracted and *z*-scored relative to the Control group. Then, two indices for each ROI were computed: i) a ‘magnitude of atrophy’ index (sum of left and right values), capturing total amount of atrophy relative to Controls, with smaller numbers indicating greater total bilateral atrophy, and ii) an ‘asymmetry of atrophy’ index (subtracting values of right from left ROI) where negative scores indicate left-lateralized atrophy, positive scores indicate right-lateralized atrophy, and scores at/around zero indicate no particularly marked lateralization of atrophy. Group differences were examined on the magnitude score but not on the asymmetry score as it does not index better/worse performance, rather, laterality of atrophy.

Complementing this analysis, we computed whole-brain voxel-level inter-subject variance in grey matter intensity to derive brain regions showing uniformly low voxel-level variance. This analysis aids interpretation of VBM correlation findings, as regions with uniformly low voxel-level variance across cases (coupled with low variation in test scores) may not emerge in VBM correlation analyses, despite their importance in explaining the cognitive-behavioural profile of patients.^30^

### Correlations of grey matter intensity with PCA-generated factor scores

Finally, VBM correlation analyses were run in the patient group to examine relationships between whole-brain grey matter intensity and performance on emergent PCA factors. A covariate-only statistical model with a positive *t*-contrast was employed with age included as a nuisance variable. We note that for our ‘Behavioural changes’ factor (Factor 1), we found multiple high-loading measures in *both* positive and negative directions; therefore, for this factor, we employed an additional correlation model with a negative *t*-contrast (with age as a nuisance variable). This step is in accordance with previous studies employing two-tailed PCA-VBM correlations specifically for factors comprising measures that load bidirectionally.^8^ Anatomical locations of statistical significance were overlaid on the MNI standard brain with maximum coordinates provided in MNI stereotaxic space. Clusters were extracted using the voxel-wise method with a strict threshold of *p*<.001 uncorrected for multiple comparisons with a cluster threshold of 50 spatially contiguous voxels to capture changes in subcortical regions that may relate to emergent factors from the PCA.

### Data availability

The ethical requirement to ensure patient confidentiality precludes public archiving of our data. Researchers who would like to access the raw data should contact the corresponding author who will liaise with the ethics committee that approved the study, and accordingly, as much data that is required to reproduce the results will be released to the individual researcher.

## Results

### Demographic, clinical, neuropsychological, and behavioural performance

BvFTD patients were significantly younger than Controls (*p*=.0009). No other significant group differences emerged for demographic factors (all *p* > .1) (Table 1; SD subgroup findings in Supplementary Results and Supplementary Table 3). Importantly, patient groups displayed comparable ages of disease onset, disease duration, and clinician-indexed disease severity (all *p*>.1). Patient groups also displayed significant general cognitive impairment (ACE-R Total) relative to Controls (*p*<.001), however, performed comparably to each other on this measure (*p*=.1). Relative to Controls, bvFTD and SD groups also displayed significant impairments on targeted measures of language, attention and working memory, verbal fluency and emotion recognition and affect selection functions (all *p*<.01) (Table 1). In addition, executive (TMT B-A) and visuospatial (ACE-R Visuospatial Total and ROCF) dysfunction was evident in bvFTD (all *p*<.05), while SD displayed significant, disproportionate deficits on single word repetition (*p*<.001).

**Table 1.**
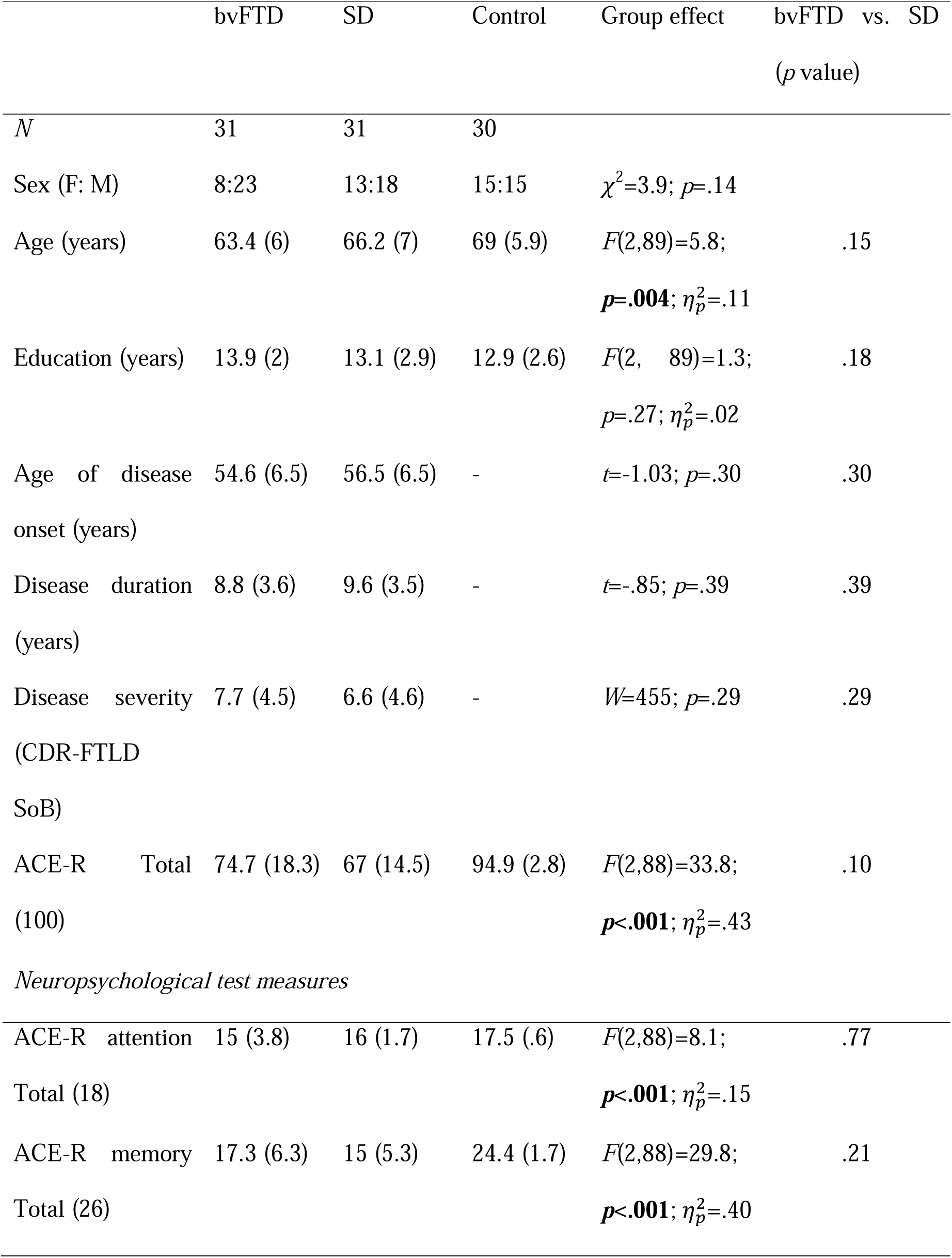
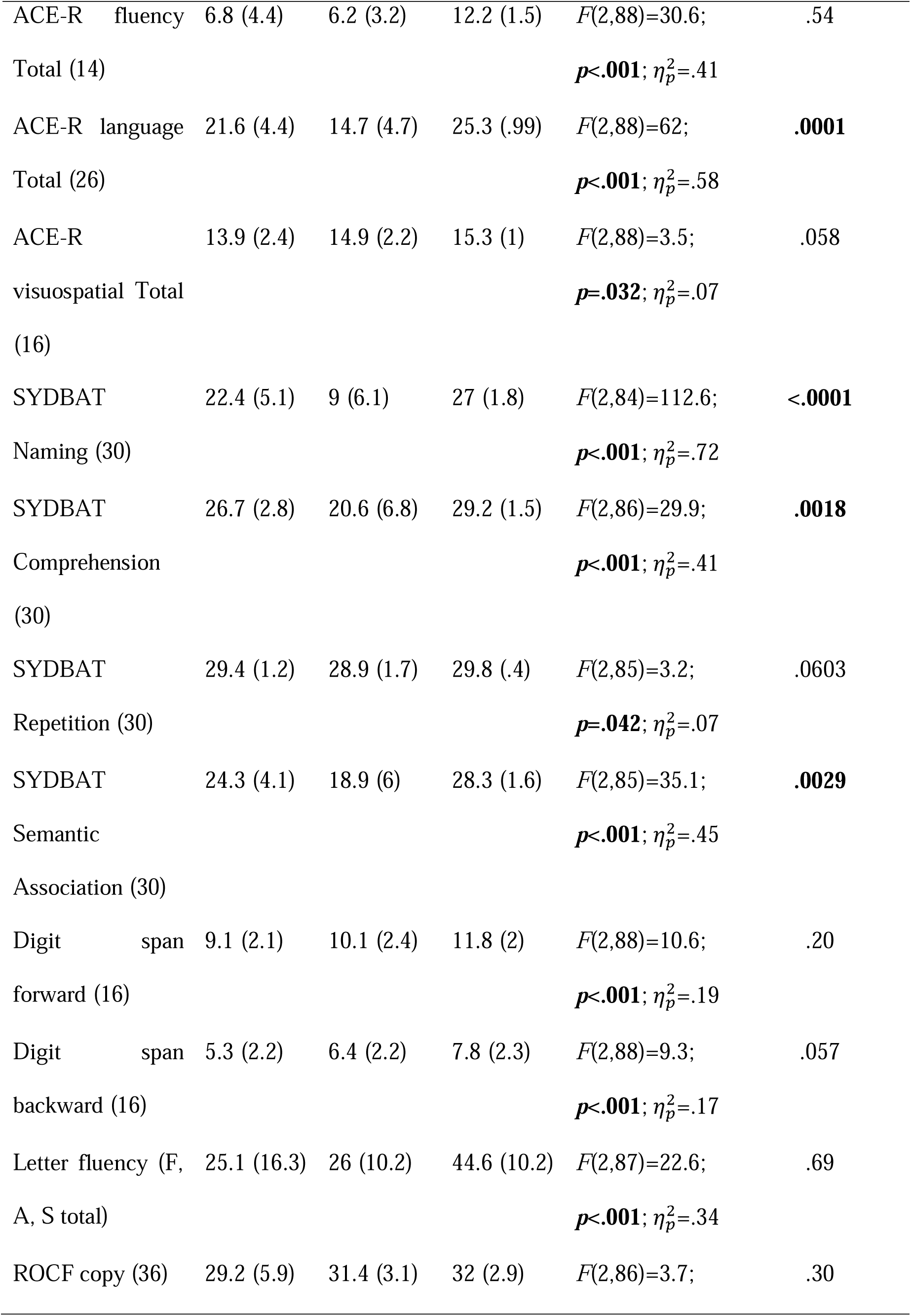
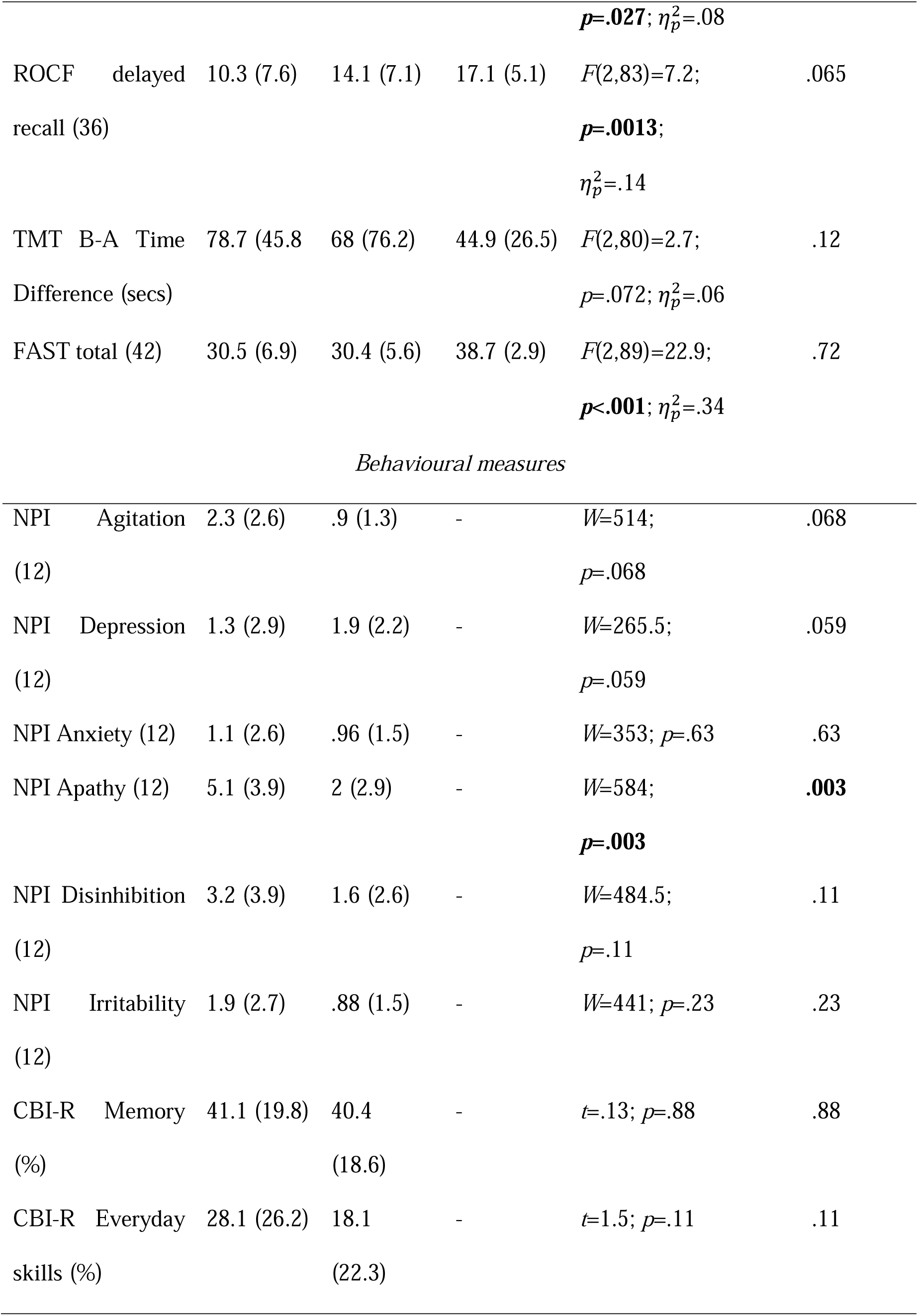
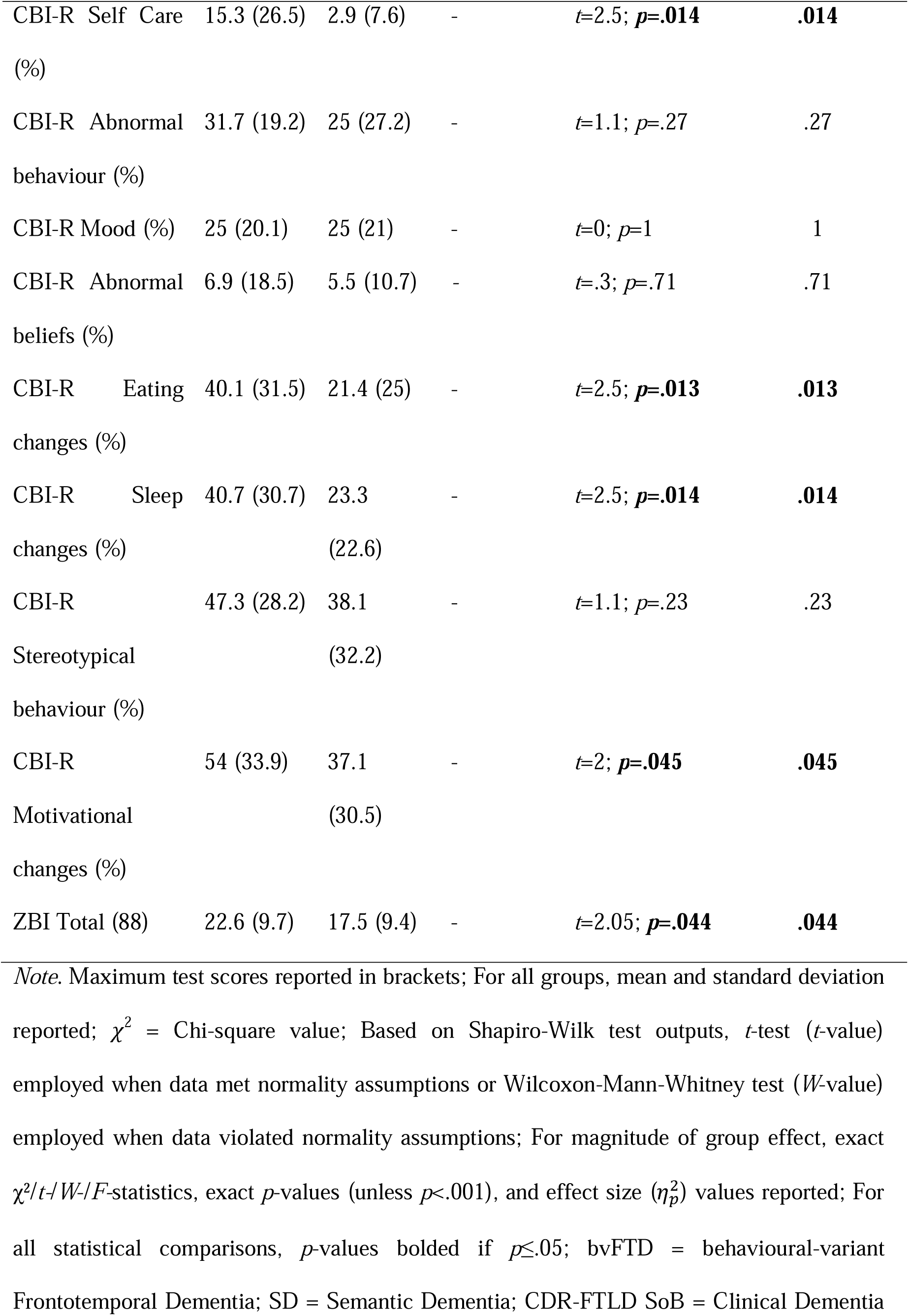
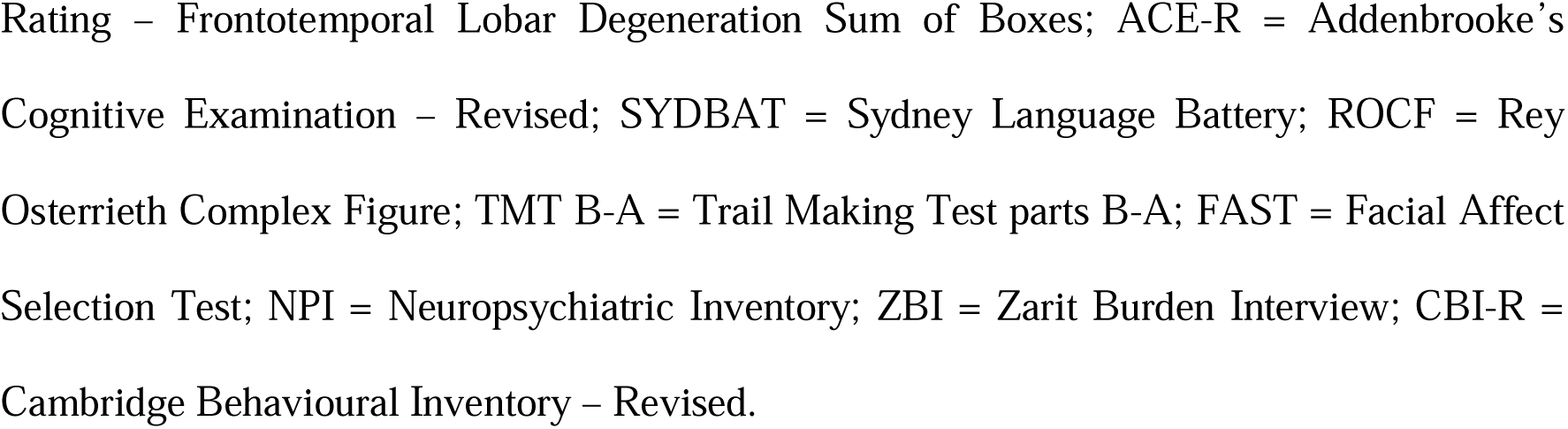
Demographic, clinical, behavioural, and neuropsychological assessment performance for all groups.

Direct comparison between bvFTD and SD groups revealed no significant differences across all cognitive domains assessed (Table 1), providing preliminary evidence, at the group-level, for pervasive overlap in cognitive dysfunction in these syndromes despite comparable disease staging. Similarly, many carer-rated behavioural disturbances were not found to differ significantly between bvFTD and SD (Table 1; all *p*>.059) however, motivational changes, self-care difficulties, eating and sleeping abnormalities were rated as more severe in bvFTD relative to SD (all *p*<.05). Carers of bvFTD patients further reported significantly greater burden relative to SD carers (*p*=.044).

### Patterns of whole brain grey matter atrophy

Next, we present results from whole brain/ROI grey matter atrophy and variance analyses as these provide a snapshot of co-occurring frontotemporal degeneration in these groups. These findings are used as a foundation to explore transdiagnostic patterns of cognitive-behavioural variation arising from degeneration of common brain systems in these syndromes.

Relative to Controls, bvFTD patients displayed reduced grey matter intensity centred on bilateral lateral and medial prefrontal cortices, insula, and OFC, extending towards temporal poles and ATL, medial temporal and striatal regions, and laterally to lateral temporal, posterior parietal, and occipital and cerebellar regions (Supplementary Table 1 and Supplementary Figure 4). Relative to Controls, SD patients displayed reduced grey matter intensity centred on the bilateral temporal poles and ATL, extending dorsally into bilateral OFC, insula and frontal poles, and medially into medial temporal and posterior parietal regions. Comparing patient groups, bvFTD patients displayed greater atrophy to bilateral prefrontal and anterior cingulate cortices compared to SD, while SD patients displayed greater atrophy to bilateral lateral and medial temporal cortices compared to the bvFTD group.

### Whole brain voxel-level inter-subject grey matter variance

Visual inspection of grey matter intensity variance maps revealed uniformly low voxel-level variance in bilateral prefrontal (OFC), insula, ATL, posterior temporal, striatal and subcortical, parietal midline, and cerebellar regions. These findings align with those from our VBM atrophy analyses as many of these regions represent sites of maximal atrophy in our bvFTD and SD group, whereas regions flanking the ‘edges’ of atrophy clusters show greater grey matter intensity variance (Supplementary Figure 4).

### Inter-subject variance in atrophy to disease-specific epicentres

#### Magnitude of atrophy

BvFTD, SD-Left, and SD-Right patients demonstrated comparable magnitude of atrophy to bilateral OFC and anterior insular cortices (all *p*>.06; Figure 1 and Supplementary Table 2). On scatterplots, this is visible as dense overlap between bvFTD and SD patients, especially for OFC integrity (Figure 1). For temporal regions, in contrast, the magnitude of ATL/temporal pole atrophy was significantly greater in SD (and in both SD subgroups) compared to bvFTD patients (all *p*<.001; Figure 1 and Supplementary Table 2).

**Figure 1.**
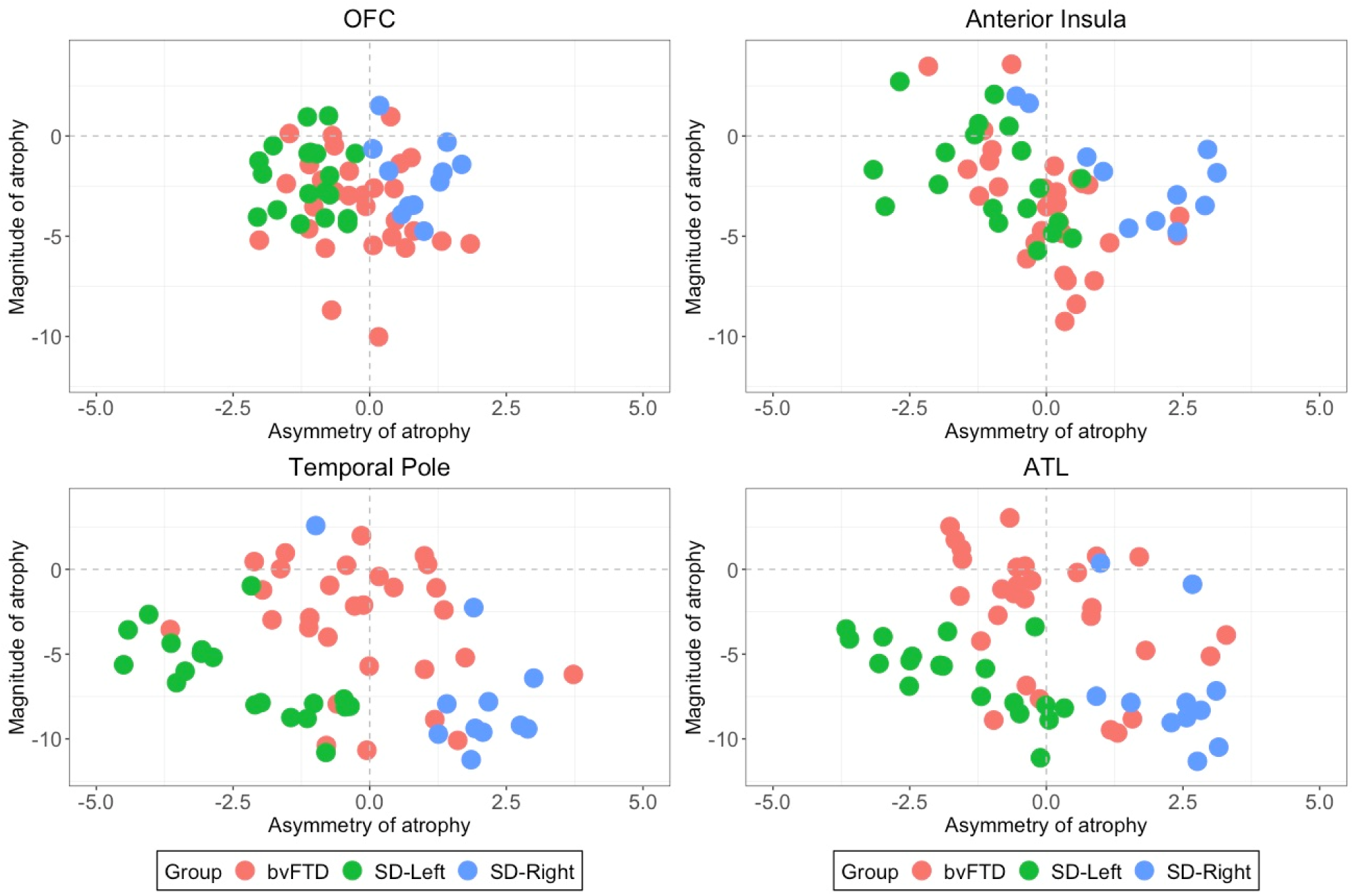
Variance in magnitude and asymmetry of atrophy in bvFTD and SD in disease-specific epicentres. All scores are *z*-scored (relative to Control group) mean intensity values for *a priori* bi-hemispheric regions of interest. The magnitude index (i.e., left+right intensity values) captures total amount of atrophy relative to Controls, and the asymmetric index (i.e., left-right score) captures asymmetry of atrophy. On the asymmetry index, negative scores suggest left-lateralized atrophy, positive scores suggest right-lateralized atrophy, and scores at/close to zero scores indicate no particularly marked lateralization of atrophy. bvFTD = behavioural variant frontotemporal dementia; SD = semantic dementia.

#### Lateralization of atrophy

For prefrontal cortex ROIs, the bvFTD group displayed evenly distributed atrophy in left and right OFC and anterior insular cortices. For temporal regions, the bvFTD group displayed relatively even bi-hemispheric atrophy in temporal poles, while most patients tended to display left-lateralized ATL involvement (Figure 1). For SD, in contrast, atrophy in both prefrontal and temporal regions was lateralised greatly to the hemisphere primarily affected in their individual SD subgroup diagnosis (Figure 1).

### Determining principal factors underlying cognitive-behavioural performance

Emergent factors and test loadings from the PCA are displayed in Figure 2 and Supplementary Table 4. The sample size was considered adequate for PCA (Kaiser-Meyer-Olkin statistic=.69). Both probabilistic and varimax-rotated PCAs converged on a solution with 8 orthogonal components with eigenvalues>1, together explaining 73.4% of the performance variance in the patient cohort (Supplementary Figures 5-6).

**Figure 2.**
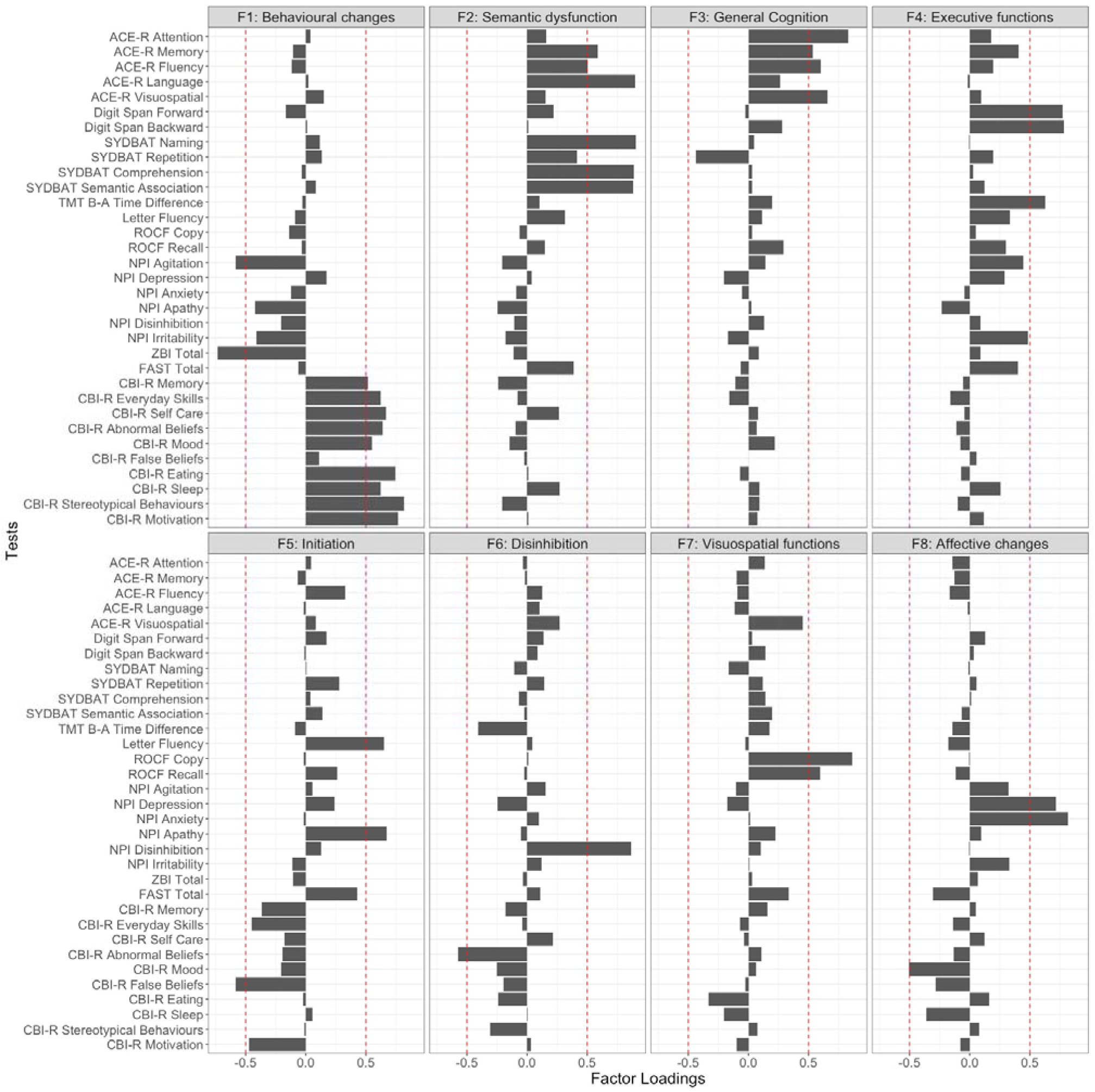
Factor loadings for neuropsychological and behavioural measures in the combined behavioural variant frontotemporal dementia and semantic dementia group (*N*=62) on a varimax-rotated 8 component PCA solution. Panel indicates emergent factors, in the order of the amount of overall variance explained. Red dashed lines representing factor loading cut-offs (>0.5 or <-.05). ACE-R = Addenbrooke’s Cognitive Examination – Revised; SYDBAT = Sydney Language Battery; TMT B-A = Trail Making Test parts B-A; ROCF = Rey-Osterrieth Complex Figure; NPI = Neuropsychiatric Inventory; ZBI = Zarit Burden Interview; FAST = Facial Affect Selection Task; CBI-R Cambridge Behavioural Inventory – Revised.

Factor 1 was labelled ‘Behavioural changes’, explained 21.5% of the overall variance, and loaded positively on measures of behavioural changes (CBI-R stereotypical and abnormal behaviour, motivation, eating, self-care and everyday skills, sleep, mood, and memory subscales) and negatively with the NPI Agitation score and overall carer burden (ZBI Total), suggesting patients with more preserved everyday behaviours (e.g., motivation, sleep, self-care, mood) were less likely to be agitated and were associated with less carer burden.

Factor 2 was titled ‘Semantic dysfunction’, captured 18.5% of the overall variance, and loaded mainly on tasks of semantic functions such as SYDBAT Naming, Comprehension, and Semantic Association subtests, and ACE-R Language, Memory, and Fluency subscales.

Factors 3 and 4 accounted for 8.5% and 6.9% of the total variance, respectively. Tests loading on Factor 3 included ACE-R Memory, Fluency, Attention, and Visuospatial subscales and was labelled ‘General cognition’. Factor 4 was named ‘Executive functions’ and included measures of sustained attention and working memory (Digit Span Forward and Backward) and overall executive performance and processing speed (TMT B-A measure).

The fifth and sixth factors respectively captured 5.4% and 4.6% of the overall variance. Factor 5 was named ‘Initiation’ and mainly loaded on Letter Fluency and NPI Apathy scores, as well as the CBI-R false beliefs measure. Factor 6 was referred to as ‘Disinhibition’ as it loaded on the NPI disinhibition and CBI-R abnormal behaviour subscales.

The seventh factor explained 3.9% of the overall variance and was labelled ‘Visuospatial functions’, loading heavily on ROCF Copy and Delayed Recall components. The eighth factor was named ‘Affective changes’, captured 3.8% of the overall variance, and included the CBI-R Mood, and NPI depression and anxiety subscales. Measures not loading heavily on any factors included the SYDBAT Repetition, FAST Total score, and the NPI irritability subscore.

### Graded overlap and differences in performance in bvFTD and SD

Individual-level performance, group-level gradations, and scatter plots for select factors are presented in Figure 3, with comparisons between bvFTD, SD, and SD-subgroups displayed in Supplementary Results and Supplementary Figures 7 and 8. Marked overlap across original diagnostic templates was noted (Figure 3A). At the group-level, bvFTD and SD displayed comparable performance on Behavioural changes (Factor 1; *t*=1.9, *p*=.058), General Cognition (Factor 3; *t*=-1.3, *p*=.18), Initiation (Factor 5; *t*=1.3, *p*=.17), Disinhibition (Factor 6; *t*=-.83, *p*=.41), and Affective changes (Factor 8; *t*=-1.3, *p*=.18) factors. In contrast, disproportionate impairments were noted in bvFTD relative to SD on the Executive (Factor 4; *t*=-2.08, *p*=.041) and Visuospatial (Factor 7; *t*=-2.05, *p*=.044) factors, whereas SD patients displayed poorer performance on the Semantic factor (Factor 2; *t*=6.9, *p* <.0001) when compared to bvFTD.

**Figure 3.**
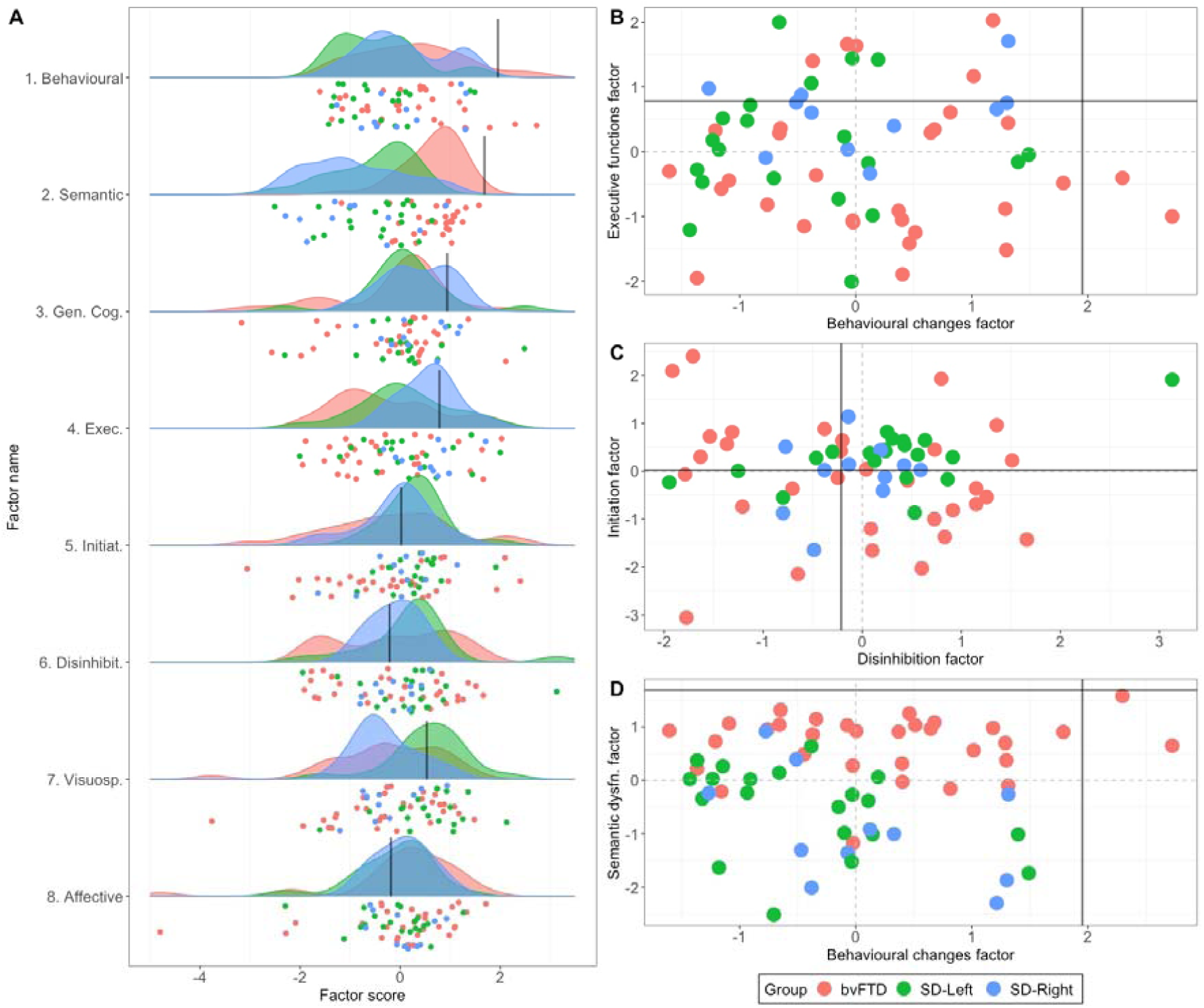
Factor scores for bvFTD and SD patients on emergent factors. **A)** Raincloud plots showcasing group-level density of distribution (cloud) and individual jittered points (rain) of performance variation across PCA-derived factors for bvFTD and SD patients. Scatter plots for select factors displaying relationships between **B)** Behavioural changes (Factor 1) with Executive function (Factor 4), **C)** Initiation (Factor 5) and Disinhibition (Factor 6) factors, and **D)** Behavioural changes (Factor 1) and Semantic dysfunction (Factor 2) factors. In all plots, black lines indicate lower bound of normality (−1.96 standard error from the mean) as estimated from the Control group (calculation detailed in Supplementary Methods). Positive scores approaching lower bound of normality indicate better performance. bvFTD = behavioural variant frontotemporal dementia; SD = semantic dementia.

### Neural correlates of principal cognitive-behavioural factors

Associations between grey matter intensity and PCA-generated factor scores in patients are displayed in Figure 4 and Supplementary Table 5. Irrespective of clinical diagnosis, Behavioural changes (Factor 1) was associated with grey matter intensity changes in left OFC, frontal pole and anterior cingulate cortex, right inferior frontal gyrus, left supramarginal gyrus and parietal operculum cortex, and bilateral posterior cingulate cortex and precuneus. Semantic dysfunction (Factor 2) was associated with grey matter intensity in bilateral ATLs and lateral temporal cortices, including bilateral temporal poles extending ventrally into temporal fusiform gyri and medially into the right medial temporal lobe (hippocampus, amygdala, putamen). Regions in the left insular cortices and Heschl’s gyrus, lateral temporal cortices (bilateral middle/inferior temporal gyri), and the left inferior parietal cortex (angular gyrus) also emerged as significant. Performance on the General cognition factor (Factor 3) was associated with left-lateralized occipital, parietal (precentral/postcentral gyrus) and subcortical (left thalamus) regions. For the Executive functions factor (Factor 4), prefrontal regions such as bilateral superior frontal and left middle/inferior frontal gyri, left insular cortex, and the left frontal pole emerged as significantly associated with performance. Initiation (Factor 5) difficulties were associated with grey matter alterations in bilateral prefrontal (frontal medial cortex, frontal poles, OFC, superior frontal gyrus, and paracingulate gyrus), bilateral subcortical (putamen, caudate, thalamus, and left pallidum and nucleus accumbens), as well as right insular cortices. Finally, performance on the Visuospatial factor (Factor 7) was associated with right ventral temporal regions including temporal/temporo-occipital fusiform and lingual gyri as well as the right posterior parahippocampal cortex. These specific regions are not areas of maximal atrophy in bvFTD and SD, but rather, flank regions of maximal atrophy and are areas of greater grey matter variance (Supplementary Figure 4). No significant clusters emerged for Disinhibition (Factor 6) or Affective changes (Factor 8) factors at *p*_unc_<.001.

**Figure 4.**
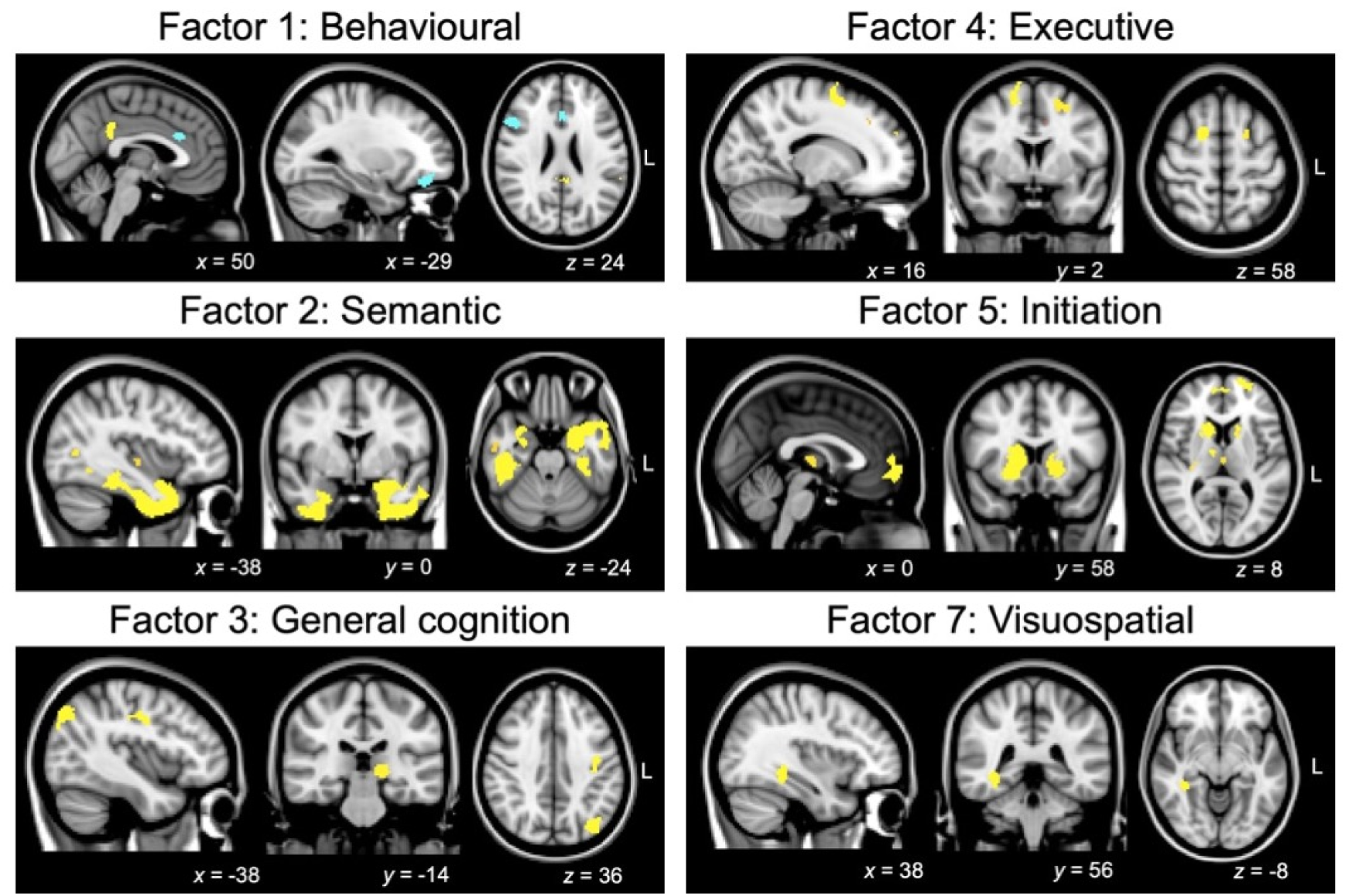
Regions of grey matter intensity correlating with PCA-generated factors in the patient cohort. All factors are derived from varimax-rotated PCA of cognitive-behavioural measures in the combined bvFTD-SD group. Coloured voxels indicate regions emerging as significant in the voxel-based morphometry analyses at a threshold of *p*<.001 uncorrected for multiple comparisons with a cluster threshold of 50 contiguous voxels (yellow denotes positive associations; blue denotes negative associations). All clusters reported at *t*=3.97 (see Supplementary Table 5 for detailed cluster information). Age was included as a nuisance covariate in the analysis. Clusters are overlaid on the Montreal Neurological Institute (MNI) standard brain with *x, y*, and *z* co-ordinates reported in MNI standard space. L = Left; bvFTD = behavioural variant frontotemporal dementia; SD = semantic dementia; PCA = Principal Component Analysis.

## Discussion

This study demonstrates marked overlap on cognitive-behavioural performance in a large group of well-characterised patients clinically labelled as bvFTD and those labelled as SD. Using PCA, we identified eight independent, separable dimensions explaining cognitive-behavioural performance variations along which clinical entities of bvFTD and SD were systematically located. Notably, bvFTD and SD showed systematic variation, undifferentiated at a group-level, across five of the emergent factors, namely Behavioural changes, General Cognition, Initiation, Disinhibition, and Affective changes. Performance on these factors reflected the differential degeneration of fronto-insular, parietal, and striatal regions. In contrast, we found evidence of discrete factors that better differentiated between the patient groups. Semantic dysfunction was, unsurprisingly, associated with SD and reflected ATL and temporal lobe atrophy, while Executive and Visuospatial dysfunction were more characteristic of bvFTD, attributable to fronto-insular and ventral temporal atrophy, respectively.

Before discussing our findings and their clinical implications in detail, it is important to pause and understand what our transdiagnostic conceptualisation of FTD (here, restricted to bvFTD and SD) represents and what it *does not* mean. Traditional approaches to exploring cognitive-behavioural variations in FTD seek out mutually exclusive diagnostic categories and subcategories. As newer symptoms and their variations are discovered, subcategories continue to grow with a continuing sequence of studies proposing new subtypes and categories falling under an ‘FTD space’. This pursuit is perfectly reasonable, provided there is sufficient intra-category homogeneity and inter-category separation that is reliably detected across studies, cohorts, and techniques. In clinical practice and across independent bodies of evidence,^8,12,32^ however, graded performance variations within and between mutually exclusive categorisations are evident. Much of this variation is closely tied to individual differences in underlying neural degeneration, pathological processes, and disease progression.^50^ Our transdiagnostic approach aids extraction of these systematic performance variations observed across patients and also maps inter-individual variations within a graded FTD space. This approach allows to preserve the well-known separation of prototypical clinical descriptions of ‘pure’ SD and bvFTD as situated in different areas within this space; yet the same model can also accommodate graded patient variations and overlapping symptom dimensions. Dimensions, however, *do not reflect* new categories of symptomatic changes or performance deficits; rather they are axes of the multidimensional space. In accordance with this view, the clinical translation of our findings is that while bvFTD and SD display marked heterogeneity across multiple cognitive-behavioural dimensions, this heterogeneity is *graded* and not absolute between syndromes, and varies at the individual-level.

Using whole-brain VBM, we validated our PCA approach by demonstrating discrete neuroanatomical signatures of emergent factors, which are in line with the extant cognitive neuroscience and neurology literature. For example, our neural correlates of Behavioural changes, Initiation, and Executive functions resonate with a large body of work implicating bilateral fronto-insular and striatal regions in the origin of these disturbances in FTD.^51-57^ In contrast, Semantic disturbances, as expected, were associated with bilateral temporopolar, ATL, and posterior portions of the ATL bordering posterior temporal/inferior parietal regions. Bilateral ATLs form trans-modal hubs of semantic processing in the brain,^5^ the earliest sites of metabolic and structural changes in SD,^58^ which progress along the ATL correlating with increasing semantic impairment in FTD.^59^ In the temporal lobe, we further found right ventral temporal regions to correlate with Visuospatial changes in FTD. These regions are important for the integration of complex visuospatial information^60^ and have typically received attention in the SD-Right literature for their potential role in mediating visuospatial and face-processing difficulties.^61^ We note, however, that these ventral temporal sites are not regions of earliest atrophy in bvFTD and SD-Left. Indeed, they showed greater inter-subject grey matter variance, therefore, possibly displayed increased sensitivity to being detected by our VBM correlation analyses. Finally, posterior parietal cortices and subcortical thalamic areas emerged as associated with performance on the General Cognition factor. Degeneration of the parietal cortex, although typically emerging in later FTD disease stages,^62^ is a key candidate for executive, general cognitive, and behavioural dysfunction, as found in investigations of the behavioural/dysexecutive variant of Alzheimer’s disease.^63,64^ On the other hand, striatal and thalamic structural degeneration are present early in the disease course in FTD^62,65,66^ and may explain a variety of goal-directed disturbances in FTD.^53,67^ Collectively, these results indicate that (i) performance gradations emerge amidst overlapping distributions of frontal and temporal atrophy across bvFTD and SD; and (ii) distinct areas of grey matter changes underpin dimensional performance along each respective factor.

It is important to consider these findings with respect to current classification practices of FTD. While the international consensus criteria for bvFTD^35^ and SD^36^ have been invaluable for refining the identification and characterisation of FTD patients worldwide, they do not acknowledge the degree of overlap between cases and the difficulty that many cases present in terms of classification. Although we have shown considerable overlap between bvFTD and SD across a range of cognitive and behavioural features, we do not by any means endorse that all such cases should be lumped together as simply FTD. Rather, we envisage that new methods to precisely identify pathological subgroups *in vivo* will enable us to better understand the pathophysiology of these cases, and ultimately move towards curative drug therapies targeted to such pathologies. Nevertheless, there are strong grounds for considering SD as a unique clinico-pathological syndrome within the FTD spectrum. Such cases typically progress slowly, are rarely genetically determined,^9^ and are strongly associated with a singular pathology (FTLD-TDP Type C pathology) which is seldom found in other cases of FTD.^68^ It should be acknowledged, however, that clinico-pathological studies have, to date, not revealed any key features predictive of pathology, beyond broad associations between SD with FTLD-TDP Type C pathology.^1^ The degree of association between SD and this specific pathology has further been shown to vary, perhaps reflecting some of the inherent overlap in clinical features with bvFTD,^69^ as highlighted by the present study. Syndromic classification in FTD is also challenging as patients present at different clinical stages depending on disease staging and progression.^12^ Transdiagnostic approaches could therefore prove critical in identifying predictive clinical features which definitively map onto pathology post-mortem. Longitudinal studies of this kind are extremely challenging and time consuming to conduct but are of the utmost importance to accelerate our understanding of the links between phenotype, clinical features, and underlying pathological drivers in bvFTD and SD.

In this vein, some limitations of our study warrant discussion. First, the majority of our patients have not yet come to autopsy, preventing us from conducting clinico-pathological explorations. We constrained our imaging analyses to explore only grey matter changes and reported our VBM correlation results at an uncorrected threshold of *p*<.001. This threshold has been suggested as far more conservative than corrections for multiple comparisons and is increasingly adopted in studies of cognition and behaviour in neurodegenerative syndromes.^70,71^ Nevertheless, it is important that future studies use larger cohorts and integrate detailed clinical, cognitive-behavioural assessments with indices of pathological, genetic, natural history, and multimodal neuroimaging (structural, functional, and diffusion imaging) approaches. This will allow to comprehensively model the multifaceted relationships between different mechanisms contributing to heterogeneity in FTD.^72^

Our findings hold a number of clinical implications. Mapping cognitive-behavioural variations to a unified FTD space affords potential opportunities to explore clinical management and symptomatic treatment options benefitting multiple disease groups rather than just one diagnostic category. Uncovering common dimensions of phenotypic change opens further possibilities to design treatment plans that harness common moderators of phenotypes to slow disease progression. By revealing the neural architecture of heterogeneous symptoms, we can consider the possibility of medical and functional restoration programmes targeting specific brain structural and functional networks. Our transdiagnostic approach also offers a fine-grained accompaniment to current categorical diagnostic approaches to capture symptomatic heterogeneity at individual, group, and neural levels. Crucially, our findings can inform future revisions of the international diagnostic criteria for bvFTD and SD to accommodate graded differences in phenotypic presentation as characteristic of both syndromes. Adopting this approach in pathologically confirmed FTD patients can further refine our understanding of the primary syndrome and likely pathologies associated with mixed cognitive presentations of these syndromes. By creating such multidimensional spaces, it may also be possible to simplify longitudinal mapping of disease evolution and to explore disease progression in prototypical cases who may progress to show ‘atypical’ symptoms. This, in turn, may improve the capacity to provide tailored management and care information for patients, their families and carers. These remain important avenues for future work.

## Supplementary material

Supplementary material is available at *Brain* online.

