## Supplementary material for "Mapping behavioural, cognitive and affective transdiagnostic dimensions in frontotemporal dementia"

**SUPPLEMENTARY METHODS**

**General and targeted neuropsychological assessment**

Participants underwent comprehensive neuropsychological testing including targeted assessments of memory, language, and executive functioning. Global cognitive functioning was assessed using the ACE-R total score and its subtests of attention and orientation, memory, verbal fluency, language, and visuospatial processing (Mioshi *et al.*, 2006). Targeted assessments of language performance included confrontational naming, single-word comprehension, single-word repetition, and semantic association subtests of the Sydney Language Battery (SYDBAT; Savage *et al.*, 2013). Each SYDBAT subscale has a maximum score of 30 with demonstrated sensitivity to differentiating profiles of language deficits in primary progressive aphasia syndromes, such as SD (Savage *et al.*, 2013). Auditory attention and working memory were assessed using forward and backward digit span tests (Wechsler, 1997). Controlled word generation and retrieval were measured using the verbal letter fluency (F, A, S) test (Strauss *et al.*, 2006). Visuo-constructional ability and visuospatial recall were indexed using the Copy and 3-minute delayed recall components of the Rey-Osterrieth Complex Figure (ROCF; Osterrieth, 1944) (max score = 36). Executive dysfunction was assessed using the time difference between parts B and A of the Trail Making Test (TMT B-A; Reitan, 1958). Finally, the total score of the Facial Affect Selection Task (FAST) component of the Emotion Selection Task (Miller *et al.*, 2012) (max score = 42), requiring participants to match a target face to one of six emotions (fear, surprise, anger, disgust, happiness, sadness), was used as a metric of emotion recognition and affect selection. It should be noted that a preliminary PCA that included each emotional component of the FAST as an independent measure revealed these components to be tightly correlated and load together on the same factor, regardless of the component solution derived. We, therefore, opted to use the total score of the FAST as a reliable index of overall affect selection performance.

**Statistical analyses**

*Converting raw scores to percentages*

For tests with maximum scores attainable and linear scoring patterns (where larger scores indicate better performance), data were converted into percentages using the formula: [(raw score / max score) * 100] (namely for the ACE-R subtests, SYDBAT subtests, digit spans, ROCF subtests, FAST, CBI components, and ZBI total). By contrast, the letter fluency test had no maximum attainable score and therefore, we used a ‘local maximum’ score (i.e., the highest verbal fluency score indicative of the best performance) as a substitute for max score. Similarly, the TMT B-A and NPI component measures followed an inverse scoring pattern (where larger scores indicate poorer performance), with the TMT B-A measure additionally characterized by no maximum attainable score. For these two measures, a two-step standardization process was adopted. First, TMT B-A and NPI component scores were converted into percentages using the ‘local maximum’ score (i.e., the highest TMT B-A/NPI score, indicative of poorest performance) using the formula: [(raw score / local maximum) * 100]. This score was subsequently subtracted from 100 to derive an index of TMT B-A/NPI performance percentage.

*Imputing missing data with Probabilistic Principal Component Analysis*

Missing data were imputed using a probabilistic PCA approach. This approach allows for the estimation of the number of underlying principal components in the entire dataset (including missing data) and uses this information to probabilistically predict missing values. As compared to list-wise exclusion of rows of missing data or imputation with central tendency values, probabilistic PCA offers improved stability and guards against overfitting of imputed data points (see Tipping and Bishop, 1999; Ilin and Raiko, 2010).

We used a *k*-fold cross-validation approach (with *k* = 4) where the dataset was first divided into three ‘training’ sets and one ‘testing’ set (as *k* = 4). Following this, a *k*-fold cross-validation probabilistic PCA with 1,000 permutations was used to estimate the component solution (*i.e.,* the number of components) that represent the underlying structure of the data. The solution with the lowest root-mean-square-error for all held-out cases (i.e., testing set cases) over 1,000 permutations was considered the most ‘stable’ solution (Ballabio, 2015). This approach was also used to select the optimal number of components for subsequent PCA on the imputed dataset. All imputed scores were visually inspected by S.R. to ensure they fit with each patient’s overall cognitive profile.

*Projecting lower bound of normality from Control data into the patient PCA-space*

All Control data were first standardized into percentages using methods detailed above. For each test, the ‘lower bound of normality’ (-1.96 standard error of the mean) was estimated, and *z*-scored using the mean and standard deviation of the respective test. The *z*-scored lower bound of normality value for each test was then multiplied with its respective component coefficient (*i.e.*, loading) from the PCA run in the patient group to receive a product. This product reflected the Control lower bound of normality for each measure included in the patient PCA. As we were interested in a single value representative of the lower bound of normality for each of the PCA-generated factors, we retained only products for tests loading highly (loadings > .5) on each factor. The retained products were summed to derive a single value reflecting the Control lower bound of normality in the patient PCA space for that specific factor. This value was then projected onto the patient PCA plot. It should be noted that Controls did not have NPI scores, therefore, this variable was excluded when computing the lower bound of normality for each factor.

*Voxel-based morphometry pre-processing*

Voxel-based morphometry (VBM) analyses were conducted on 3D T_1_-weighted structural MR images to identify voxelwise changes in grey matter intensity across groups using the FSL-VBM toolbox (Ashburner and Friston, 2000), from the FMRIB software package (<https://fsl.fmrib.ox.ac.uk/fsl/fslwiki/fslvbm/index.html>) (Smith *et al.*, 2004). The VBM pre-processing analysis comprised a standard pipeline involving brain-extraction of structural MR images using FMRIB’s Brain Extraction Tool (BET: Smith, 2002). Next, brain-extracted images were subject to tissue segmentation using FMRIB’s Automatic Segmentation Tool (FAST: Zhang *et al.*, 2001). Then, brain-extracted tissue segmented images were aligned to the Montreal Neurological Institute standard space (MNI52) using a b-spline representation of the registration warp field (Rueckert *et al.*, 1999) implemented in the FMRIB non-linear registration approach (FNIRT: Andersson *et al.*, 2007a; Andersson *et al.*, 2007b). All resultant images were then collated into a study-specific template, to which a nonlinear re-registration of native grey matter images was performed. The registered partial volume maps were then modulated by dividing by the Jacobian of the warp field to correct for local expansion or contraction. The final, modulated and segmented images were smoothed using an isotropic Gaussian kernel with a sigma of 3 mm.

To investigate grey matter intensity differences between groups, an unbiased whole-brain general linear model was used, incorporating permutation-based non-parametric testing with 5,000 permutations per contrast (Nichols and Holmes, 2002). Regression models with separate directional contrasts (*i.e.*, *t*-tests) were used to assess differences in cortical grey matter intensities between bvFTD and Controls, and between SD and Controls groups, with age included as a nuisance variable. Clusters were then extracted using the Threshold-Free Cluster Enhancement method using a threshold of *p* < .01 corrected for Family -Wise Error (FWE) with a cluster threshold of 100 spatially contiguous voxels.

*Region-of-interest selection for magnitude and asymmetry of atrophy in disease-specific epicenters*

For each participant, mean grey matter intensities were extracted using binarized masks derived either from the Harvard-Oxford Cortical Atlas in FSLview (for OFC and temporal pole), meta-analytic based atlases (ATL; Rice *et al.*, 2015), or reliable multimodal parcellation atlases (anterior insula; Kelly *et al.*, 2012). As the ATLs envelopes the temporal poles, the ATL mask used here excluded the temporal poles so as to capture the gradation of atrophy along the temporal cortex in SD (Supplementary Figure 3).

**SUPPLEMENTARY RESULTS**

**Demographic and clinical variables in SD-Left and SD-Right subgroups**

Supplementary Table 1 displays demographic, clinical, and neuropsychological test performance in bvFTD, SD-Left, and SD-Right groups. The bvFTD group was significantly younger than the SD-Left group (*p* = .048). No other significant differences emerged between participant groups on demographic factors such as sex distribution or years of education (both *p* > .1). Turning to clinical variables, bvFTD, SD-Left, and SD-Right groups demonstrated comparable ages of disease onset, disease duration, clinician-indexed disease severity (CDR-FTLD SoB), as well as general cognitive status, as examined on the ACE-R Total score (all *p* values > .1).

**Neuropsychological test performance in SD-Left and SD-Right subgroups**

Neuropsychological testing performance for bvFTD, SD-Left and SD-Right groups are displayed in Supplementary Table 1. Relative to the Control group, both SD-Left and SD-Right groups demonstrated significantly poor performance on global measures of attention, memory, fluency, and language performance (as measured by subtests of the ACE-R) as well as targeted assessments of confrontational naming, single word comprehension, semantic association, letter fluency, and emotion recognition and affect selection (all *p* values < .05). In comparison to Controls, SD-Left patients additionally displayed impairments on auditory attention and working memory measures (both *p* < .05) while SD-Right patients exhibited poor visuospatial recall (*p* < .05). Direct comparison between bvFTD and SD groups indicated that both SD subgroups performed significantly poorer than the bvFTD group on primary language tasks (ACE-R Language, SYDBAT naming and comprehension) (all *p* values < .01; Supplementary Table 1). In contrast, the bvFTD group performed significantly poorer than the SD-Left group on visuospatial cognitive measures, including the ACE-R Visuospatial Total and ROCF recall (both *p* < .05), and worse than the SD-Right group on a measure of working memory (Digit Span Backward) (*p* < .01). No significant differences emerged between SD-Left and SD-Right subgroups on any neuropsychological test measures (all *p* values > .07; Supplementary Table 1).

**Behavioural disturbances in SD-Left and SD-Right subgroups**

Direct comparisons between patient groups revealed greater carer-reported apathy, self-care difficulties, abnormal behaviour, eating, sleeping and motivational changes in the bvFTD group relative to the SD-Left group (all *p* values < .05). No significant differences emerged when investigating behavioural disturbances between bvFTD and SD-Right, or SD-Left and SD-Right groups (all *p* values > .05) (Supplementary Table 1).

**Group differences on principal cognitive-behavioural factors between bvFTD, SD-Left, and SD-Right groups**

Group comparisons on emergent factor scores revealed that the SD-Left group scored significantly lower than the bvFTD group on the Behavioural changes factor (Factor 1; *p* = .03). On the Semantic dysfunction factor (Factor 2), both SD groups performed significantly poorer than the bvFTD group (all *p* values < .001), but comparably to each other (*p* > .1). On the Executive factor (Factor 4), the bvFTD group performed significantly worse than the SD-Right group (*p* = .007). On the Visuospatial factor (Factor 7), both bvFTD and SD-Right patients performed poorer than the SD-Left group (both *p* values < .01), with no significant differences between bvFTD and SD-Right groups (*p* > .1). No other significant differences emerged.

**Supplementary Table 1.** Voxel-based morphometry results showing regions of significant grey matter intensity reductions in patient groups relative to Controls and between patient groups

| Regions | Side | Number of voxels | Peak MNI co-ordinate | | | *t*-value |
| --- | --- | --- | --- | --- | --- | --- |
|  |  |  | *x* | *y* | *z* |  |
| *bvFTD < Controls* | | | | | | |
| Superior/middle/inferior frontal gyrus, medial frontal cortex, anterior/paracingulate cortex, insular cortex, frontal pole, orbitofrontal cortex, extending ventrally towards temporal poles, anterior temporal lobes, superior/middle/inferior temporal gyrus, planum temporale, Heschl’s gyrus, hippocampus, parahippocampal cortex, amygdala, thalamus, caudate, nucleus accumbens, putamen, extending postero-dorsally into supramarginal/angular gyrus, precuneus, precentral/postcentral gyrus, parietal opercular cortex, posterior cingulate cortex, lateral occipital cortex and left occipital pole, bilateral lingual gyrus, temporo-occipital gyrus, temporal fusiform cortex and cerebellum | Bilateral | 62,219 | 42 | -50 | -58 | 3.75 |
| Intracalcarine cortex, lingual gyrus, occipital pole, occipital fusiform, and inferior lateral occipital cortex | Right | 1,299 | 14 | -88 | 8 | 2.9 |
| *SD < Controls* | | | | | | |
| Temporal fusiform cortex, temporal pole, inferior/middle/superior temporal gyrus extending antero-dorsally towards bilateral frontal opercular cortex, inferior frontal gyrus, orbitofrontal cortex, insular cortex, left frontal pole, extending postero-ventrally into bilateral parahippocampal gyrus, hippocampus, amygdala, putamen, left thalamus, bilateral temporo-occipital fusiform cortex, left Heschl’s gyrus and planum temporale, right lingual gyrus, extending postero-dorsally into left supramarginal/angular gyrus, left precentral/postcentral gyrus, left precuneus, left superior parietal lobule, and left central/parietal opercular cortex | Bilateral | 30,304 | -26 | -12 | -50 | 3.75 |
| Superior frontal gyrus, precentral/postcentral gyrus, precuneus and superior parietal lobule | Right | 1,094 | 32 | -30 | 62 | 3.17 |
| Supramarginal gyrus, postcentral gyrus and parietal opercular cortex | Right | 242 | 42 | -30 | 34 | 2.66 |
| Insular cortex extending to central and parietal opercular cortex, | Right | 227 | 32 | -10 | 16 | 2.63 |
| Frontal pole and middle frontal gyrus | Right | 184 | 26 | 42 | 20 | 2.78 |
| *bvFTD < SD* | | | | | | |
| Frontal pole, orbitofrontal cortex, medial frontal cortex, subcallosal cortex, inferior/superior frontal gyrus, frontal opercular cortex, insular cortex, anterior/paracingulate gyrus, extending into right precentral gyrus and right middle frontal gyrus | Bilateral | 8,359 | -42 | 24 | -10 | 3.75 |
| Left paracingulate gyrus, left superior frontal gyrus, left anterior cingulate gyrus, right paracingulate gyrus, right anterior cingulate gyrus, right juxtapositional lobule cortex | Bilateral | 270 | -2 | 20 | 44 | 2.71 |
| *SD < bvFTD* | | | | | | |
| Temporal fusiform cortex, temporal pole, parahippocampal gyrus, lingual gyrus, temporal occipital fusiform cortex, amygdala, putamen, planum polare,  Inferior/middle/superior temporal gyrus, insular cortex | Left | 3,763 | -28 | 10 | -46 | 3.53 |
| Temporal fusiform cortex, inferior temporal gyrus, parahippocampal gyrus and temporal pole | Right | 251 | 40 | -12 | -36 | 2.64 |

*Note.* MRI data were available for 29 bvFTD and 30 SD [19 SD-Left, 11 SD-Right] patients. Clusters presented emerged as significant in the VBM analyses using Threshold-Free Cluster Enhancement at *p* < .01 corrected for Family-Wise Error with a strict cluster threshold of 100 contiguous voxels. Age was included as a nuisance variable in the analyses. MNI = Montreal Neurological Institute; bvFTD = behavioural variant frontotemporal dementia; SD = semantic dementia.

**Supplementary Table 2**. Magnitude and asymmetry of atrophy in disease-specific epicenters in bvFTD and SD groups.

|  | bvFTD | SD-Left | SD-Right | Magnitude of group effect | Post-hoc results (direction of effect) |
| --- | --- | --- | --- | --- | --- |
| Left+right OFC | -3.4 (2.4) | -2.1 (1.7) | -2.0 (1.8) | *F* (2, 56) = 2.9; *p* = .060; $\eta_{p}^{2}$ = .09 | - |
| Left-right OFC | -.1 (.8) | -1.1 (.55) | .8 (.5) |  |  |
| Left+right anterior insula | -3.4 (3) | -2.0 (2.4) | -1.9 (2.3) | *F* (2, 56) = 2.07; *p* = .13; $\eta_{p}^{2}$ = .06 | - |
| Left-right anterior insula | .04 (1) | -.9 (1.1) | 1.6 (1.2) |  |  |
| Left+right ATL | -2.5 (3.7) | -6.2 (2.1) | -7.1 (3.6) | *F* (2, 56) = 11.3; ***p* < .001**; $\eta_{p}^{2}$ = .28 | bvFTD > SD-Left, SD-Right |
| Left-right ATL | .03 (1.3) | -1.5 (1.3) | 2.3 (.7) |  |  |
| Left+right temporal pole | -3.2 (3.6) | -6.3 (2.4) | -7.3 (4) | *F* (2, 56) = 7.8; ***p* < .001**; $\eta_{p}^{2}$ = .21 | bvFTD > SD-Left, SD-Right |
| Left-right temporal pole | -.14 (1.4) | -2.3 (1.3) | 1.8 (1) |  |  |

*Note*. All scores are *z*-scores standardized relative to mean and standard deviation of the Control group. No statistical comparisons were conducted for asymmetry analyses (i.e., left-right) as the outcome index does not measure better or worse performance but rather the laterality of atrophy. On the asymmetry index, negative scores suggested left-lateralized atrophy, positive scores suggested right-lateralized atrophy, and scores at/close to zero scores indicated bilateral atrophy of relatively equal magnitude. MRI data were available for 29 bvFTD and 30 SD [19 SD-Left, 11 SD-Right] patients. bvFTD = behavioural variant frontotemporal dementia; SD = semantic dementia; OFC = orbitofrontal cortex; ATL = anterior temporal lobe (excluding temporal pole).

**Supplementary Table 3**. Demographic, clinical, and behavior and general neuropsychological assessment performance for bvFTD, SD-Left and SD-Right groups.

|  | bvFTD | SD-Left | SD-Right | Group effect | bvFTD vs. SD-Left (*p* value) | bvFTD vs. SD-Right (*p* value) | SD-Left vs. SD-Right (*p* value) |
| --- | --- | --- | --- | --- | --- | --- | --- |
| *N* | 31 | 20 | 11 |  |  |  |  |
| Sex (F: M) | 8:23 | 8:12 | 5:6 | $\chi$^2^ = 1.8; *p* = .38 |  |  |  |
| Age (years) | 63.4 (6) | 67.7 (6.5) | 63.5 (7.4) | *F* (2, 59) = 2.8; *p* = .064; $\eta_{p}^{2}$ = .08 | **.048** | .08 | .18 |
| Education (years) | 13.9 (2) | 13.3 (2.8) | 12.7 (3) | *F* (2, 59) = 1.09; *p* = .34; $\eta_{p}^{2}$ = .03 | .34 | .24 | .71 |
| Age of disease onset (years) | 54.6 (6.5) | 57.4 (7.9) | 54.9 (6.5) | *F* (2, 58) = 1.01; *p* = .37; $\eta_{p}^{2}$ = .03 | .41 | .76 | .68 |
| Disease severity (CDR-FTLD SoB) | 7.7 (4.5) | 5.9 (4.2) | 7.7 (5.2) | *F* (2, 53) = .91; *p* = .4; $\eta_{p}^{2}$ = .03 | .23 | .83 | .43 |
| ACE-R Total (100) | 74.7 (18.3) | 66.7 (13.9) | 67.4 (16.3) | *F* (2, 58) = 1.6; *p* = .19; $\eta_{p}^{2}$ = .05 | .09 | .24 | .82 |
| *Neuropsychological test measures* | | | | | | | |
| ACE-R attention total (18) | 15 (3.8) | 16.1 (1.5) | 15.8 (2.1) | *F* (2, 58) = .86; *p* = .42; $\eta_{p}^{2}$ = .02 | .86 | .81 | .83 |
| ACE-R memory total (26) | 17.3 (6.3) | 14.5 (5) | 16 (5.8) | *F* (2, 58) = 1.3; *p* = .26; $\eta_{p}^{2}$ = .04 | .14 | .59 | .58 |
| ACE-R fluency total (14) | 6.8 (4.4) | 6.2 (3.1) | 6.2 (3.5) | *F* (2, 58) = .14; *p* = .86; $\eta_{p}^{2}$ < .01 | .68 | .72 | .87 |
| ACE-R language total (26) | 21.6 (4.4) | 14.8 (4.5) | 14.6 (5.3) | *F* (2, 58) = 17.07; ***p* < .001**; $\eta_{p}^{2}$ = .37 | **< .0001** | **.0007** | .87 |
| ACE-R visuospatial total (16) | 13.9 (2.4) | 15 (2.6) | 14.7 (1.3) | *F* (2, 58) = 1.3; *p* = .27; $\eta_{p}^{2}$ = .04 | **.026** | .64 | .26 |
| SYDBAT Naming (30) | 22.4 (5.1) | 8.2 (5) | 10.4 (7.6) | *F* (2, 55) = 41.3; ***p* < .001**; $\eta_{p}^{2}$ = .60 | **< .0001** | **.0002** | .54 |
| SYDBAT Comprehension (30) | 26.7 (2.8) | 22.5 (5.4) | 17.1 (8) | *F* (2, 57) = 15.2; ***p* < .001**; $\eta_{p}^{2}$ = .34 | **.006** | **.0007** | .35 |
| SYDBAT Repetition (30) | 29.4 (1.2) | 28.9 (2.1) | 29 (1) | *F* (2, 56) = .63; *p* = .53; $\eta_{p}^{2}$ = .02 | .33 | .08 | .38 |
| SYDBAT Semantic (30) | 24.3 (4.1) | 20.2 (5.8) | 16.4 (5.8) | *F* (2, 56) = 10.5; ***p* = .0001**; $\eta_{p}^{2}$ = .27 | **.01** | **.0002** | .15 |
| Digit span forward (16) | 9.1 (2.1) | 10.1 (2.5) | 10.2 (2.4) | *F* (2, 58) = 1.3; *p* = .27; $\eta_{p}^{2}$ = .04 | .27 | .32 | .82 |
| Digit span backward (16) | 5.3 (2.2) | 6 (2.4) | 7.2 (1.4) | *F* (2, 58) = 2.7; *p* = .07; $\eta_{p}^{2}$ = .08 | .29 | **.009** | .13 |
| Letter fluency (F, A, S total) | 25.1 (16.3) | 27.7 (9.9) | 23 (10.3) | *F* (2, 58) = .46; *p* = .63; $\eta_{p}^{2}$ = .01 | .227 | .86 | .39 |
| ROCF copy (36) | 29.2 (5.9) | 32.1 (2.9) | 30.3 (3.3) | *F* (2, 57) = 2.2; *p* = .11; $\eta_{p}^{2}$ = .07 | .13 | .83 | .31 |
| ROCF delayed recall (36) | 10.3 (7.6) | 15.8 (7.2) | 10.9 (5.9) | *F* (2, 54) = 3.4; ***p* = .038**; $\eta_{p}^{2}$ = .11 | **.01** | .79 | .11 |
| TMT B-A Time Difference (secs) | 78.7 (45.8 | 78.9 (91) | 46.1 (20.8) | *F* (2, 51) = 1.06; *p* = .35; $\eta_{p}^{2}$ = .03 | .35 | .07 | .37 |
| FAST total (42) | 30.5 (6.9) | 32.2 (4.5) | 27.2 (6.2) | *F* (2, 59) = 2.27; *p* < .11; $\eta_{p}^{2}$ = .07 | .58 | .17 | .07 |
| *Behavioural measures* | | | | | | | |
| NPI Agitation (12) | 2.3 (2.6) | .9 (1.3) | .9 (1.2) | *F* (2, 54) = 3; *p* = .055; $\eta_{p}^{2}$ = .1 | .16 | .22 | .86 |
| NPI Depression (12) | 1.3 (2.9) | 2.2 (2.5) | 1.2 (1.4) | *F* (2, 51) = .78; *p* = .46; $\eta_{p}^{2}$ = .02 | .07 | .45 | .6 |
| NPI Anxiety (12) | 1.1 (2.6) | 1 (1.6) | .9 (1.4) | *F* (2, 52) = .05; *p* = .95; $\eta_{p}^{2}$ < .001 | .69 | .75 | .85 |
| NPI Apathy (12) | 5.1 (3.9) | 1.2 (2) | 3.2 (3.7) | *F* (2, 54) = 6.7; ***p* = .002**; $\eta_{p}^{2}$ = .19 | **.001** | .28 | .21 |
| NPI Disinhibition (12) | 3.2 (3.9) | 1.4 (2.9) | 1.8 (2.1) | *F* (2, 52) = 1.7; *p* = .18; $\eta_{p}^{2}$ = .06 | .13 | .52 | .61 |
| NPI Irritability (12) | 1.9 (2.7) | 1.1 (1.8) | .5 (.7) | *F* (2, 52) = 1.7; *p* = .18; $\eta_{p}^{2}$ = .06 | .45 | .37 | .77 |
| ZBI Total (88) | 22.6 (9.7) | 17.2 (9.7) | 17.9 (9.2) | *F* (2, 55) = 2; *p* = .13; $\eta_{p}^{2}$ = .07 | .11 | .36 | .71 |
| CBI-R Memory (%) | 41.1 (19.8) | 40 (17.5) | 41.1 (21.3) | *F* (2, 58) = .02; *p* = .97; $\eta_{p}^{2}$ < .001 | .83 | .86 | .83 |
| CBI-R Everyday skills (%) | 28.1 (26.2) | 16.3 (19) | 21.3 (27.8) | *F* (2, 58) = 1.4; *p* = .25; $\eta_{p}^{2}$ = .04 | .21 | .44 | .79 |
| CBI-R Self Care (%) | 15.3 (26.5) | 1.3 (5.7) | 5.6 (9.8) | *F* (2, 58) = 3.3; ***p* = .041**; $\eta_{p}^{2}$ = .1 | **.0052** | .52 | .17 |
| CBI-R Abnormal behaviour (%) | 31.7 (19.2) | 20.6 (26.8) | 32.5 (27.6) | *F* (2, 58) = 1.5; *p* = .22; $\eta_{p}^{2}$ = .05 | **.031** | .77 | .17 |
| CBI-R Mood (%) | 25 (20.1) | 21.7 (19.3) | 30.6 (23.4) | *F* (2, 58) = .66; *p* = .51; $\eta_{p}^{2}$ = .02 | .6 | .58 | .35 |
| CBI-R Abnormal beliefs (%) | 6.9 (18.5) | 3.9 (9.3) | 8.3 (12.9) | *F* (2, 58) = .35; *p* = .7; $\eta_{p}^{2}$ = .01 | .81 | .22 | .21 |
| CBI-R Eating changes (%) | 40.1 (31.5) | 14.1 (20.8) | 34 (27.5) | *F* (2, 58) = 5.1; ***p* = .008**; $\eta_{p}^{2}$ = .15 | **.0019** | .71 | .052 |
| CBI-R Sleep changes (%) | 40.7 (30.7) | 17.7 (19.2) | 32.9 (25.7) | *F* (2, 58) = 4.3; ***p* = .017**; $\eta_{p}^{2}$ = .12 | **.0096** | .62 | .16 |
| CBI-R Stereotypical behaviour (%) | 47.3 (28.2) | 31.5 (31.2) | 49.4 (32) | *F* (2, 58) = 1.9; *p* = .14; $\eta_{p}^{2}$ = .06 | .064 | .85 | .15 |
| CBI-R Motivational changes (%) | 54 (33.9) | 29.1 (28.9) | 50.9 (29.5) | *F* (2, 58) = 3.8; ***p* = .028**; $\eta_{p}^{2}$ = .11 | **.013** | .85 | .076 |
| ZBI Total (88) | 22.6 (9.7) | 17.2 (9.4) | 17.9 (9.2) | *F* (2, 55) = 2.09; *p* = .13; $\eta_{p}^{2}$ = .13 | .11 | .36 | .71 |

*Note*. Maximum test scores reported in brackets; For all groups, mean and standard deviation reported; $\chi$^2^ = Chi-square value; For magnitude of group effect, exact χ²/*F-*statistics, exact *p*-values (unless *p* < .001), and effect size ($\eta_{p}^{2}$) values reported; For all statistical comparisons, *p*-values bolded if *p* ≤ .05; bvFTD = behavioural variant of Frontotemporal dementia; SD-Left = Semantic Dementia presenting with predominant left temporal atrophy; SD-Right = Semantic Dementia presenting with predominant right temporal atrophy; CDR-FTLD SoB = Clinical Dementia Rating – Frontotemporal Lobar Degeneration Sum of Boxes; ACE-R = Addenbrooke’s Cognitive Examination – Revised; SYDBAT = Sydney Language Battery; ROCF = Rey Osterrieth Complex Figure; TMT B-A = Trail Making Test parts B – A; FAST = Facial Affect Selection Test; NPI = Neuropsychiatric Inventory; ZBI = Zarit Burden Interview; CBI-R = Cambridge Behavioural Inventory – Revised.

**Supplementary Table 4**. Factor loadings for neuropsychological and behavioural assessments on the omnibus varimax-rotated Principal Component Analysis solution.

|  | **Factor 1**  Behavioural changes | **Factor 2**  Semantic dysfunction | **Factor 3**  General cognition | **Factor 4**  Executive functions | **Factor 5**  Initiation | **Factor 6**  Disinhibition | **Factor 7**  Visuospatial functions | **Factor 8**  Affective changes |
| --- | --- | --- | --- | --- | --- | --- | --- | --- |
| CBI-R Stereotypical Behaviour | **.82** | -.20 | .09 | -.10 | -.01 | -.31 | .07 | .08 |
| CBI-R Motivation | **.77** | .01 | .07 | .12 | -.47 | .03 | -.10 | -.08 |
| CBI-R Eating | **.74** | .01 | -.07 | -.07 | -.02 | -.24 | -.33 | .16 |
| CBI-R Self Care | **.67** | .26 | .08 | -.05 | -.18 | .22 | -.03 | .12 |
| CBI-R Abnormal Behaviour | **.64** | -.10 | .07 | -.11 | -.19 | **-.57** | .11 | -.13 |
| CBI-R Everyday skills | **.62** | -.08 | -.16 | -.16 | -.45 | -.04 | -.07 | -.14 |
| CBI-R Sleep | **.62** | .27 | .09 | .26 | .06 | .00 | -.21 | -.36 |
| CBI-R Mood | **.55** | -.15 | .22 | -.08 | -.21 | -.25 | .07 | **-.50** |
| CBI-R Memory | **.52** | -.24 | -.11 | -.05 | -.37 | -.18 | .16 | .05 |
| NPI Agitation | **-.58** | -.21 | .14 | .44 | .05 | .16 | -.11 | .33 |
| ZBI Total | **-.73** | -.11 | .08 | .09 | -.10 | -.04 | .03 | .07 |
| SYDBAT Naming | .12 | **.91** | .05 | .00 | .00 | -.11 | -.17 | -.01 |
| ACE-R Language Total | .02 | **.90** | .26 | -.02 | -.02 | .10 | -.11 | -.01 |
| SYDBAT Comprehension | -.03 | **.89** | .03 | .03 | .04 | -.07 | .14 | .01 |
| SYDBAT Semantic Association | .08 | **.88** | .03 | .12 | .14 | -.02 | .20 | -.07 |
| ACE-R Memory Total | -.10 | **.59** | **.53** | .41 | -.07 | -.02 | -.10 | -.13 |
| ACE-R Fluency Total | -.11 | **.50** | **.60** | .20 | .33 | .13 | -.09 | -.17 |
| ACE-R Attention Total | .04 | .16 | **.83** | .18 | .05 | -.04 | .14 | -.14 |
| ACE-R Visuospatial Total | .15 | .15 | **.66** | .10 | .08 | .27 | .45 | .00 |
| Digit Span Forward | .01 | .01 | .28 | **.78** | -.01 | .09 | .14 | .03 |
| Digit Span Forward | -.16 | .22 | -.03 | **.77** | .17 | .14 | .03 | .13 |
| TMT B-A Time Difference | -.03 | .10 | .20 | **.63** | -.09 | -.41 | .18 | -.15 |
| NPI Apathy | -.42 | -.25 | .03 | -.23 | **.67** | -.05 | .23 | .10 |
| Letter Fluency (FAS Total) | -.09 | .32 | .11 | .33 | **.65** | .04 | -.02 | -.17 |
| CBI-R False Beliefs | .11 | -.03 | .00 | .06 | **-.58** | -.20 | -.03 | -.29 |
| NPI Disinhibition | -.21 | -.11 | .13 | .09 | .13 | **.86** | .10 | -.01 |
| ROCF Copy | -.14 | -.06 | .03 | .05 | -.02 | .01 | **.86** | -.01 |
| ROCF Recall | -.03 | .15 | .29 | .30 | .26 | -.02 | **.60** | -.11 |
| NPI Depression | .17 | .04 | -.20 | .29 | .24 | -.24 | -.18 | **.72** |
| NPI Anxiety | -.12 | -.09 | -.06 | -.04 | -.02 | .10 | .01 | **.81** |
| SYDBAT Repetition | .13 | .41 | -.44 | .20 | .28 | .14 | .12 | .06 |
| FAST Total | -.06 | .39 | -.06 | .40 | .43 | .11 | .33 | -.30 |
| NPI Irritability | -.41 | -.18 | -.17 | .48 | -.11 | .12 | .01 | .33 |

*Note*. Measures that heavily load on each factor (loadings > .5 or < .-.5) are indicated in bold. Only scores for bvFTD and SD patients were entered into the Principal Component Analysis. bvFTD = behavioural-variant Frontotemporal Dementia; SD = Semantic Dementia; ACE-R = Addenbrooke’s Cognitive Examination – Revised; SYDBAT = Sydney Language Battery; ROCF = Rey Osterrieth Complex Figure; TMT B-A = Trail Making Test parts B – A; FAST = Facial Affect Selection Test; NPI = Neuropsychiatric Inventory; ZBI = Zarit Burden Interview; CBI-R = Cambridge Behavioural Inventory – Revised.

**Supplementary Table 5**. Whole brain voxel-based morphometry results showing regions of grey matter intensity that correlate with PCA-generated factors in the combined bvFTD and SD cohort.

| Regions | Side | Number of voxels | Peak MNI co-ordinates | | |
| --- | --- | --- | --- | --- | --- |
|  |  |  | *x* | *y* | *z* |
| *Factor 1 – Behavioural changes* |  |  |  |  |  |
| **Positive contrast** |  |  |  |  |  |
| Posterior cingulate cortex and precuneus | Bilateral | 138 | 0 | -44 | 36 |
| Supramarginal gyrus, and parietal operculum cortex | Left | 97 | -60 | -36 | 28 |
| **Negative contrast** |  |  |  |  |  |
| Orbitofrontal cortex, frontal pole | Left | 165 | -28 | 38 | -20 |
| Inferior frontal gyrus | Right | 109 | 54 | 16 | 20 |
| Anterior cingulate cortex | Left | 62 | 0 | 22 | 24 |
| *Factor 2 – Semantic dysfunction* |  |  |  |  |  |
| Temporal pole, parahippocampal gyrus, temporal/temporo-occipital fusiform cortex, inferior temporal gyrus, and lingual gyrus | Left | 3,842 | -24 | 4 | -48 |
| Temporal/temporo-occipital/occipital fusiform cortex, temporal pole, parahippocampal gyrus, hippocampus, amygdala, putamen, inferior/middle temporal gyrus, and inferior lateral occipital cortex | Right | 2,410 | 32 | -6 | -50 |
| Temporo-occipital fusiform cortex, inferior/middle temporal gyrus, angular gyrus, and lateral occipital cortex | Left | 278 | -40 | -56 | -10 |
| Superior/middle/inferior temporal gyrus | Left | 198 | -60 | -22 | -20 |
| Insular cortex, Heschl’s gyrus, and putamen | Left | 86 | -34 | -14 | -4 |
| Middle/inferior temporal gyrus | Right | 63 | 56 | -10 | -24 |
| *Factor 3 – General cognition* |  |  |  |  |  |
| Superior lateral occipital cortex | Left | 260 | -44 | -82 | 28 |
| Precentral and postcentral gyrus | Left | 107 | -44 | -6 | 30 |
| Thalamus | Left | 104 | -12 | -28 | -4 |
| *Factor 4 – Executive functions* |  |  |  |  |  |
| Superior frontal gyrus, precentral gyrus, and supplementary motor area | Right | 146 | 18 | 2 | 54 |
| Central opercular and insular cortex | Left | 124 | -42 | -12 | 12 |
| Superior/middle frontal gyrus | Left | 98 | -26 | 2 | 52 |
| Middle/inferior frontal gyrus (pars triangularis & pars opercularis), and frontal pole | Left | 56 | -40 | 28 | 14 |
| *Factor 5 - Initiation* |  |  |  |  |  |
| Frontal medial cortex, frontal pole, paracingulate gyrus, superior frontal gyrus, and left paracingulate gyrus | Bilateral | 1,237 | 10 | 42 | -14 |
| Orbitofrontal cortex, putamen, pallidum, caudate, and thalamus | Right | 873 | 22 | 10 | -14 |
| Putamen, caudate, and nucleus accumbens | Left | 305 | -18 | 12 | -8 |
| Frontal pole, superior frontal gyrus, and paracingulate gyrus | Right | 155 | 18 | 40 | 34 |
| Orbitofrontal cortex and frontal pole | Left | 106 | -36 | 34 | -10 |
| Superior & inferior lateral occipital cortex | Right | 91 | 40 | -72 | 14 |
| Putamen, insular cortex, Heschl's gyrus, and planum temporale | Right | 71 | 28 | -20 | 8 |
| Thalamus | Bilateral | 51 | 2 | -16 | -2 |
| *Factor 6 - Disinhibition* |  |  |  |  |  |
| No significant clusters |  |  |  |  |  |
| *Factor 7 – Visuospatial functions* |  |  |  |  |  |
| Temporal/temporo-occipital fusiform cortex, lingual gyrus, and posterior parahippocampal gyrus | Right | 96 | 36 | -42 | -12 |
| *Factor 8 – Affective changes* |  |  |  |  |  |
| No significant clusters |  |  |  |  |  |

*Note*. MRI data were available for 29 bvFTD and 30 SD [19 SD-Left, 11 SD-Right] patients. Clusters presented emerged statistically significant in the VBM analysis at a threshold of *p*<.001 uncorrected for multiple comparisons with a cluster threshold of 50 contiguous voxels. For all clusters, *t*-values=3.97. Age was included as a nuisance covariate in the analysis. MNI = Montreal Neurological Institute; bvFTD = behavioural variant frontotemporal dementia; SD = semantic dementia.

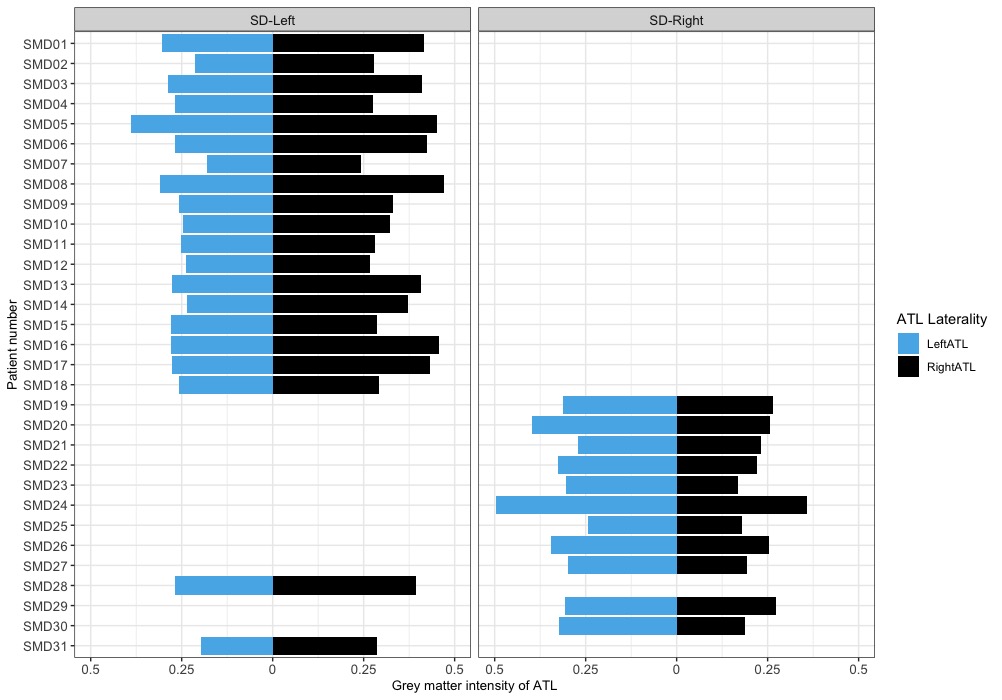

**Supplementary Figure 1.** Left and Right Anterior Temporal Lobe volumes of all semantic dementia patients included in this study (*N* = 31). Grey matter intensities of bilateral anterior temporal lobes were extracted using a binarized mask of the anterior temporal lobes from the Harvard-Oxford cortical and subcortical atlas integrated into FSLview. SD = semantic dementia; ATL = Anterior Temporal Lobe.

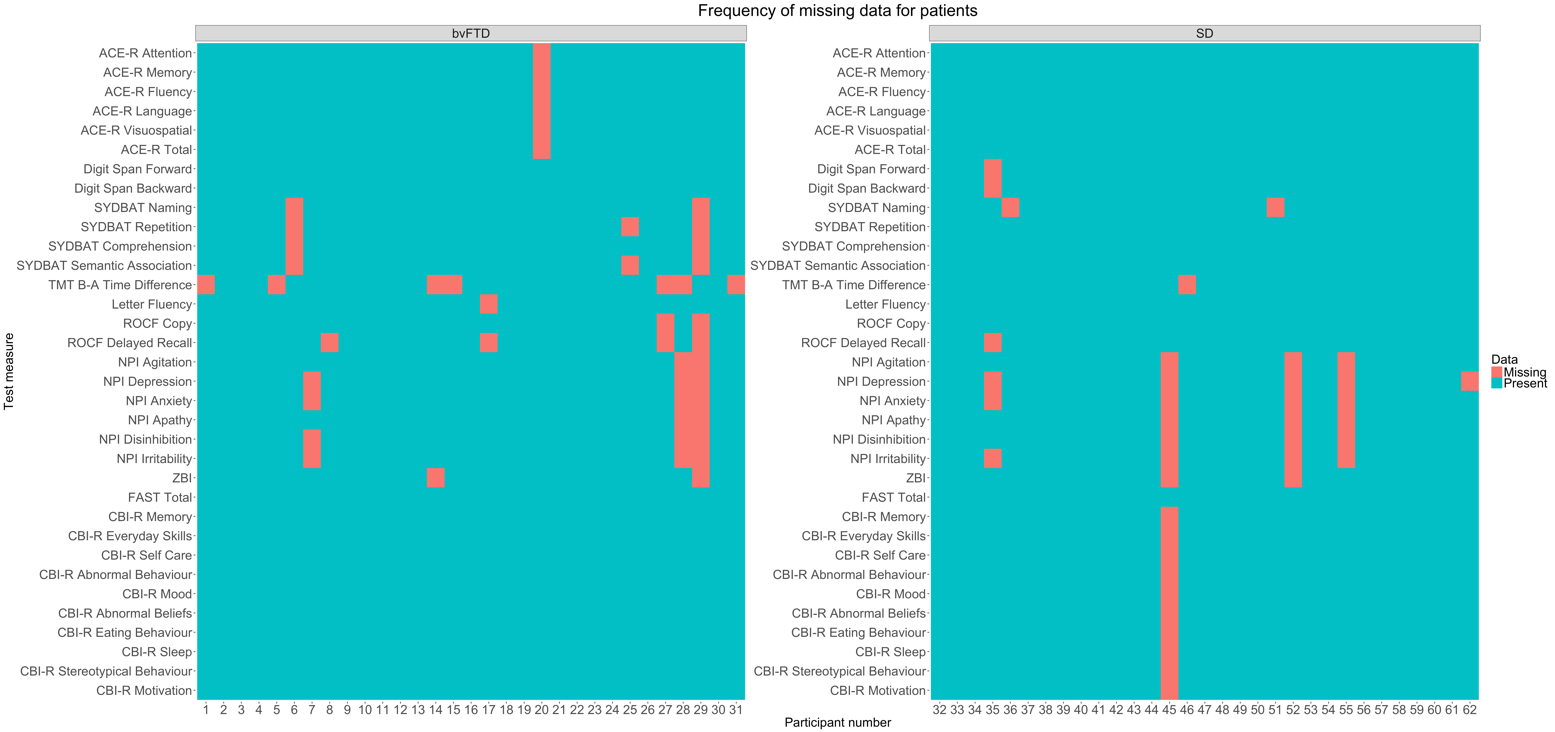

**Supplementary Figure 2**. Frequency of missing data (overall 4.17%, with 4.55% missing for bvFTD and 3.79% missing for SD; all missing data indicated as orange tiles) on neuropsychological and behavioural measures for bvFTD and SD patients. bvFTD = behavioural-variant frontotemporal dementia; SD = semantic dementia; ACE-R = Addenbrooke’s Cognitive Examination – Revised; SYDBAT = Sydney Language Battery; TMT B-A = Trail Making Test parts B – A; ROCF = Rey-Osterrieth Complex Figure; NPI = Neuropsychiatric Inventory; FAST = Facial Affect Selection Task; ZBI = Zarit Burden Interview; CBI-R Cambridge Behavioural Inventory – Revised.

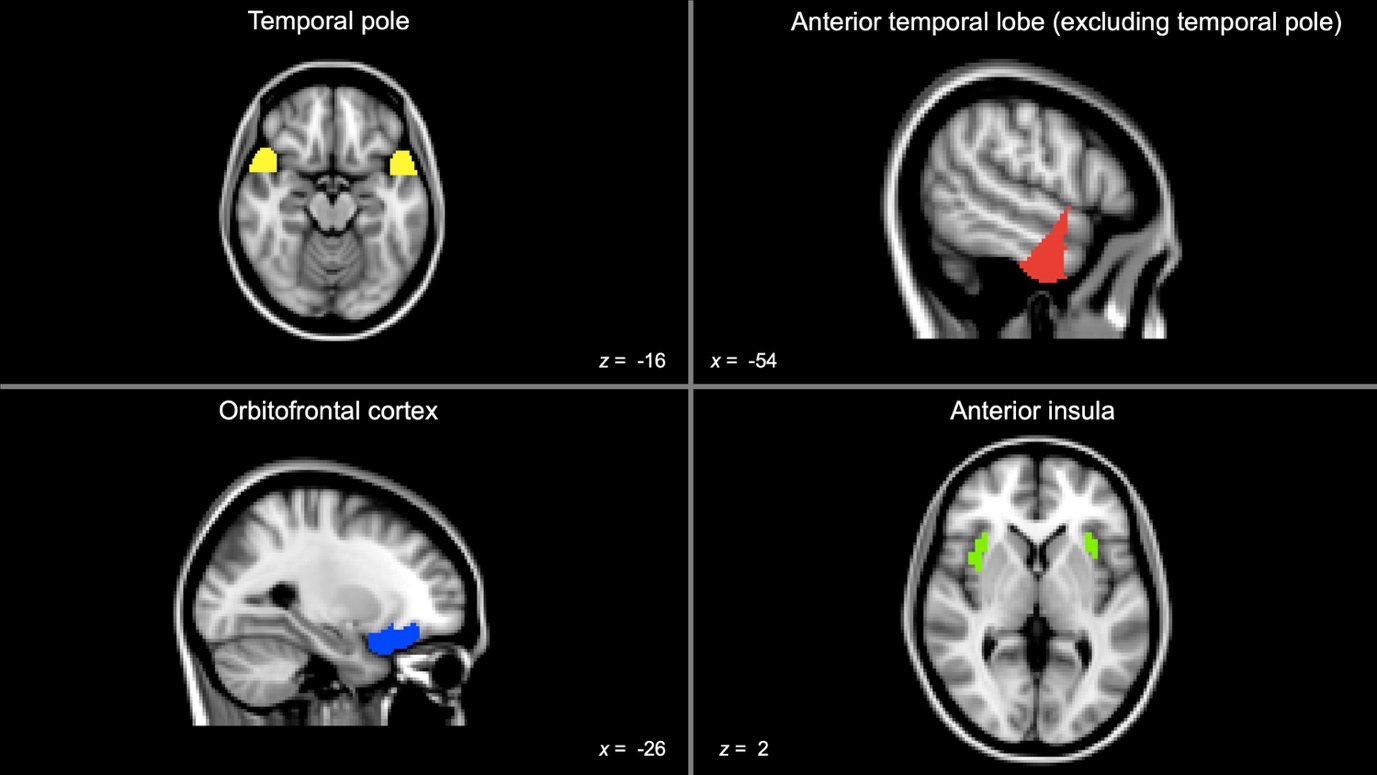

**Supplementary Figure 3.** Anatomical locations of binarized masks for four regions-of-interest for computing asymmetry of atrophy in bvFTD and SD patients. Bilateral masks for the temporal pole and orbitofrontal cortex were created using the Harvard-Oxford cortical atlas. For the anterior temporal lobe (ATL) mask employed here, a whole bilateral anterior temporal lobe mask was first derived from an activation likelihood estimation meta-analysis of functional neuroimaging studies of the ATL (Rice *et al.*, 2015), following which the binarized temporal pole mask image was subtracted from the binarized ATL mask image to derive a bilateral ATL mask excluding the temporal poles). The bilateral anterior insula mask was derived from a multimodal parcellation atlas of the insula (Kelly *et al.*, 2012). All co-ordinates are reported in Montreal Neurological Institute (MNI) space. bvFTD = behavioural variant frontotemporal dementia; SD = semantic dementia.

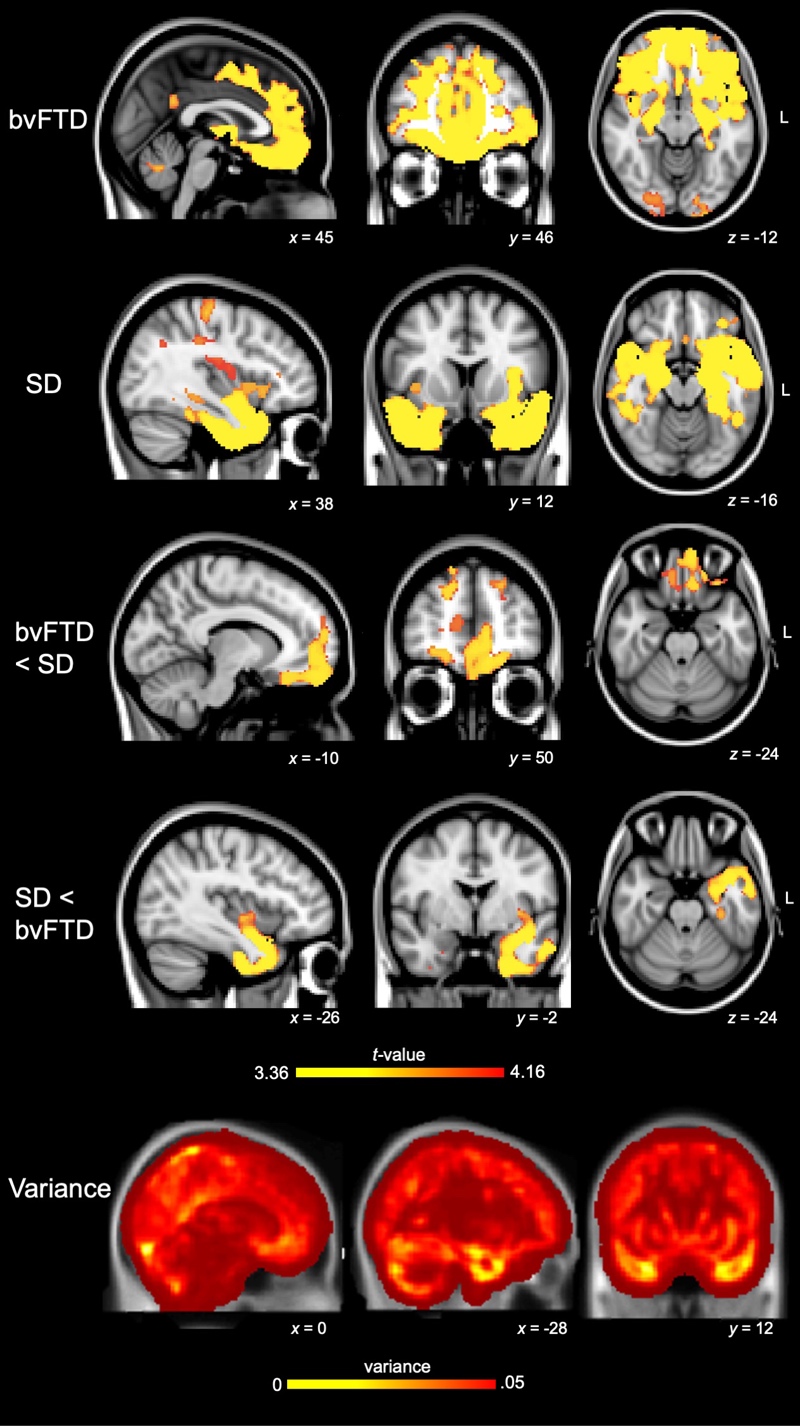

**Supplementary Figure 4.** Voxel-based morphometry (VBM) analysis of whole brain grey matter atrophy. First two panels indicate regions of significant grey matter intensity reduction in bvFTD compared to Controls and SD compared to Controls, respectively. Panel 3 and 4 indicate regions of significant grey matter intensity reduction in bvFTD compared to SD and SD compared to bvFTD, respectively. In these panels, coloured voxels indicate regions that emerged significant in the VBM analysis at *p*<.01 corrected for Family-Wise Error (FWE) with a cluster threshold of 100 spatially contiguous voxels. Age was included as a nuisance variable in this analysis. The fifth panel indicates voxel-wise variance in grey matter intensity in both patient groups compared to Controls. All clusters are overlaid on the MNI standard brain with *x*, *y*, and *z* co-ordinates reported in MNI space. bvFTD = behavioural variant frontotemporal dementia; SD = semantic dementia.

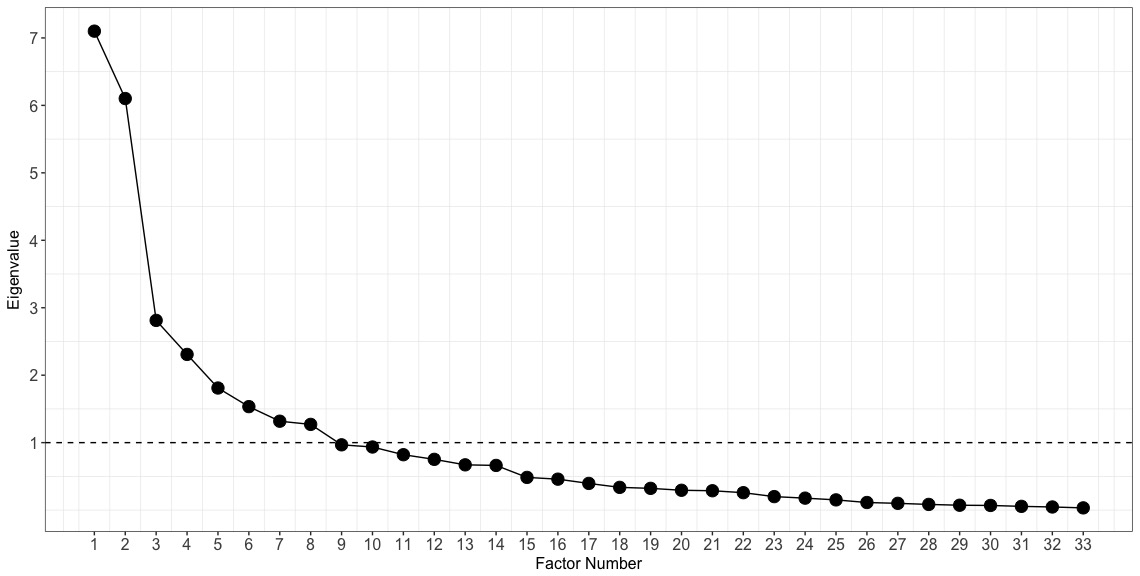

**Supplementary Figure 5**. Eigenvalues for each factor derived from PCA solution on neuropsychological and behavioural measures in bvFTD and SD patients. PCA = Principal Component Analysis; bvFTD = behavioural-variant frontotemporal dementia; SD = semantic dementia.

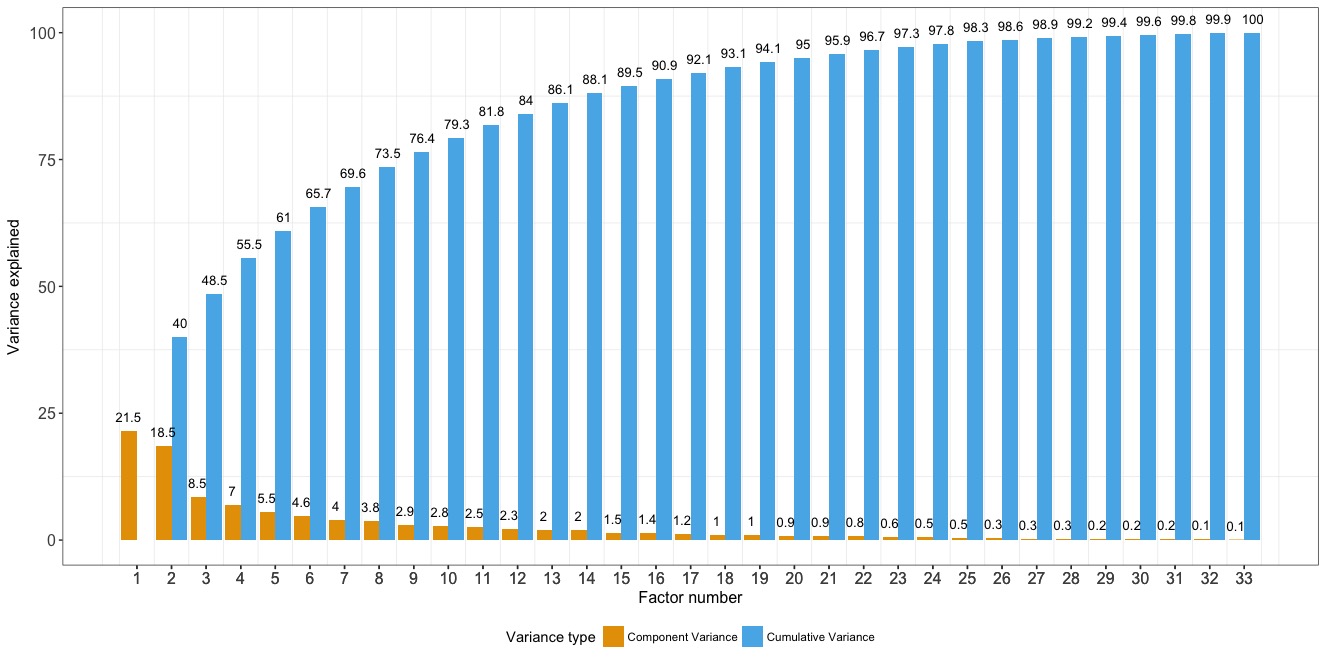

**Supplementary Figure 6**. Component-specific variance explained (yellow) and cumulative variance explained (blue) derived from the PCA solution on neuropsychological test measures in bvFTD and SD patients (with explained variance amount indicated on top of each bar). PCA = Principal Component Analysis; bvFTD = behavioural-variant frontotemporal dementia; SD = semantic dementia.

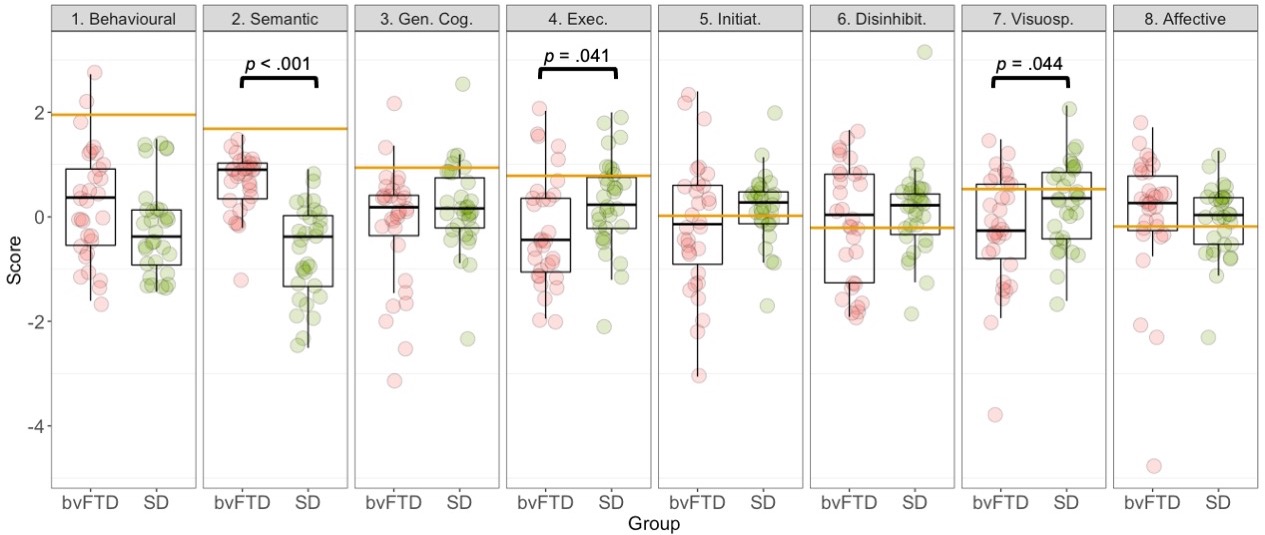

**Supplementary Figure 7.** Group differences between patients on emergent PCA factors. Statistical comparisons run using *t*-tests with *p*-values indicated. Gold lines indicate lower bound of normality (-1.96 standard error from the mean) as estimated from the Control group (calculation detailed in Supplementary Methods). bvFTD = behavioural variant frontotemporal dementia; SD = semantic dementia; PCA = Principal Component Analysis; Gen. Cog. = General Cognition factor; Exec. = Executive factor; Initiat. = Initiation factor; Disinhibit. = Disinhibition factor; Visuosp. = Visuospatial factor.

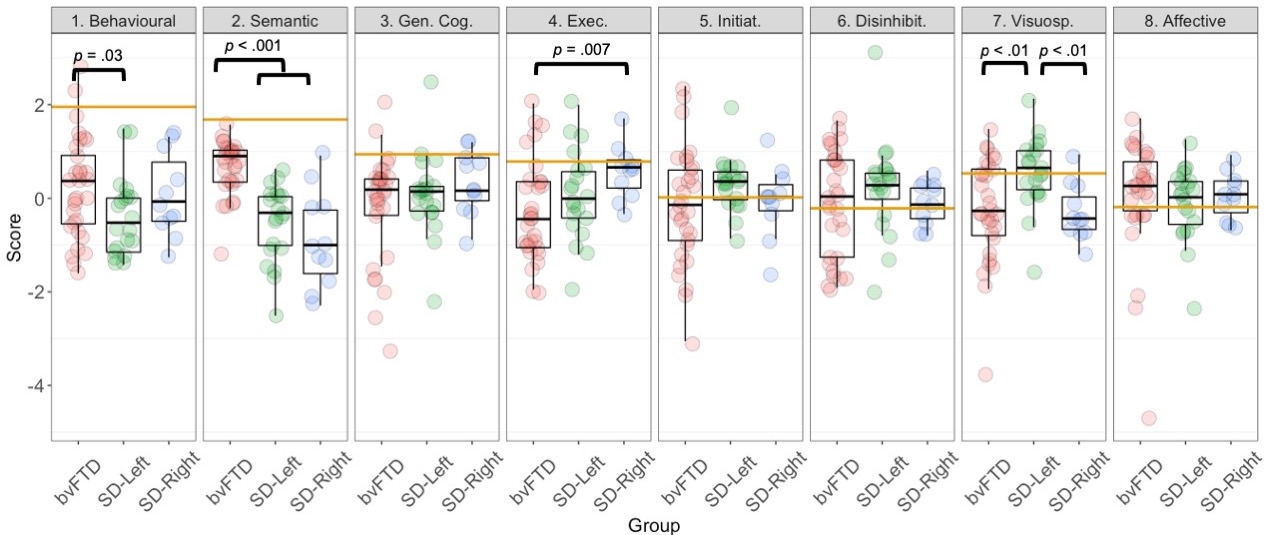

**Supplementary Figure 8.** Group differences between bvFTD, SD-Left, and SD-Right patients on emergent factors from the PCA. Statistical comparisons run using ANOVA with post-hoc comparisons using Sidak corrections. All *p*-values emerging significant in post-hoc comparisons are indicated. Gold horizontal lines indicate Control lower bound of normality score (i.e., -1.96 standard error of the mean from Control performance – see Supplementary Methods for calculation details). bvFTD = behavioural variant frontotemporal dementia; SD = semantic dementia; PCA = Principal Component Analysis; Gen. Cog. = General Cognition factor; Exec. = Executive factor; Initiat. = Initiation factor; Disinhibit. = Disinhibition factor; Visuosp. = Visuospatial factor.
